# Multimodal atlas of human atherosclerosis links granular vascular cell states to coronary artery disease risk

**DOI:** 10.64898/2026.05.24.26353986

**Authors:** Jose Verdezoto Mosquera, Ivory Tang, Maria Murach, Gaëlle Auguste, Aditi Kodali, Patrick Hart, Douglas M. Shaw, Minghong Li, Adam W. Turner, Chani J. Hodonsky, Natalia M. Dworak, Ana Karina de Oliveira, Katia Sol-Church, Teresa Jhee, Kirsten I.M. van der Sijs, Shaunak S. Adkar, Ryan B. Choi, Francesca Vacante, Joseph C. Wu, Paul Cheng, Chiara Giannarelli, Nicholas J. Leeper, Aloke V. Finn, Johan L.M. Björkegren, Jason C. Kovacic, Arif Yurdagul, Sander W. van der Laan, Clint L. Miller

**Author notes:** Correspondence: Clint L. Miller, PhD, Department of Genome Sciences University of Virginia, PO Box 800717, Charlottesville, VA 22908 USA. These authors contributed equally (Jose Verdezoto Mosquera, Ivory Tang, Maria Murach).

## Abstract

Advances in single-cell and spatial assays have revolutionized the scale and resolution of molecular tissue profiling. Here we present MetaPlaq, a multimodal atlas of human atherosclerotic arterial beds comprising over a million cells across single-cell transcriptomics, epigenomics and high-resolution spatial expression assays. We map granular cell states and disease-relevant transcriptional programs within the native tissue context of coronary arteries. Furthermore, we map cardiovascular GWAS signals to smooth muscle cells (SMCs) and endothelial cells (ECs) and uncover the *cis*-regulatory architecture governing their phenotypic transitions. Our comprehensive epigenomic reference allowed us to build cell-specific enhancer-gene link maps and multimodal gene regulatory networks (GRNs) underlying disease-relevant states such as osteogenic SMCs and ECs undergoing mesenchymal transition. We also integrate SMC and EC disease-associated gene sets with GRNs to nominate key transcription factors such as PRRX1, BNC2 and ELK3 regulating atherosclerosis-relevant transcriptional programs. Finally, we layer single-cell and spatial modalities to fine-map GWAS variants with improved cell and anatomical context. We highlight candidate cell-specific regulatory mechanisms at less characterized CAD loci, including *FGD5* and *MCF2L* in ECs. Together, this atlas represents an important step towards fully interpreting genetic risk loci and informing new therapeutic strategies for cardiovascular disease.

## Introduction

Atherosclerotic vascular diseases, such as coronary and carotid artery disease, remain leading causes of death worldwide^1^. Plaque build-up inside arterial walls often progresses to end-stage clinical events such as myocardial infarction (MI) or stroke^2^. This process involves a complex interplay of vascular and immune cell states along a continuum, which contribute to residual cardiovascular risk^3^. In response to injury of the inner vessel wall, contractile smooth muscle cells (SMCs) and endothelial cells (ECs) adopt a mesenchymal state, whereas ECs also acquire inflammatory or pro-angiogenic properties during early and advanced atherosclerosis^4,5^.

Genome-wide association studies (GWAS) have identified >300 loci associated with CAD, yet the causal cell states and regulatory mechanisms remain mostly unresolved^6,7^. We and others have shown that SMCs and ECs contribute the majority of the heritable disease risk^8–10^, highlighting the need for cell state-resolved maps of gene regulation in disease-relevant compartments.

Meta-analyses of single-cell transcriptomic datasets have expanded the catalog of vascular and immune cell states in atherosclerosis and enabled detection of rare and transitional populations linked to plaque progression^9,11,12^. However, transcriptomic studies alone provided limited insight into the spatial context of these states in native lesions. While recent spatial transcriptomics studies of human coronary and carotid arteries have begun to localize disease-associated programs, they lack resolution to define granular vascular cell states ^11,13,14^. Furthermore, transcriptomics does not directly capture the *cis*-regulatory programs driving disease-associated states. This remains a critical gap, as most GWAS variants reside in non-coding regions of the genome and likely impact disease risk through regulation of gene expression^15^.

Single-cell assays of transposase accessible chromatin (ATAC-seq)^16^ in human atherosclerotic arteries have begun to define the *cis*-regulatory architecture in vascular and immune cells^8,17^loci. However, the inherent sparsity of individual epigenomic datasets limits inference of gene regulatory networks (GRN) in less abundant disease-relevant states such as osteochondrogenic SMCs and endothelial-to-mesenchymal transition (EndoMT) ECs, and estimates of their contribution to disease heritability. In addition, robust evaluation of disease progression across multiple ancestries requires integration of large and diverse cohorts with multiple single-cell modalities, which remains a computational challenge in highly heterogeneous atherosclerotic tissues.

Here we integrate public single-cell transcriptomic datasets across four different arterial beds, with newly generated single-nucleus epigenomic, and high-resolution spatial transcriptomic profiles from human coronary arteries, to build an expanded atlas of human atherosclerosis. By profiling ∼1 million cells and nuclei across 254 samples and single cell/spatial modalities, we define more than 50 vascular and immune cell states and localize them within their anatomical context. We confirm the disease relevance of SMCs and ECs, characterize their state-specific transcriptional and cis-regulatory programs, and elucidate molecular changes associated with lesion progression. Through integration of chromatin accessibility profiles, we resolve enhancer landscapes in fibromyocytes, fibrochondrocytes, and EndoMT ECs, enabling inference of enhancer-driven gene regulatory networks (eGRNs), and refine the cell-specific interpretation of CAD risk loci. We nominate new transcription factor regulators, CAD effector genes and candidate causal variants implicated in EC states. Together, this atlas provides a powerful framework for mapping candidate causal regulatory mechanisms in atherosclerosis and for prioritizing targets linked to residual cardiovascular disease risk.

## Results

### Single cell transcriptomic map of human atherosclerosis

To generate a multimodal single-cell atlas of human atherosclerosis, we integrated in-house/public scRNA-seq and newly generated snATAC-seq datasets, yielding 361,551 transcriptomic and chromatin accessibility profiles from 216 samples across two modalities **(Fig. 1, Supplementary Table 1).** We also generated high-resolution Visium HD spatial transcriptomic data from 38 segments across 32 individuals **(Fig. 1)**. To build the annotated atlas, we harmonized transcriptomic data using our prior scRNA meta-analysis^9^ as a reference to train a single-cell variational inference (scVI) model. We then used the scArches reference mapping approach proposed by Lotfollahi et al^18^ to fine-tune reference weights with 11 new datasets and generate batch-corrected embeddings **(Fig. 2a)**. We performed detailed benchmarking showing improved balance between batch mixing and biological signal conservation relative to standalone scVI and widely-used linear models^19–22^ **(Supplementary Fig. 1d-1h).** Integration with scArches, followed by a second round of low-quality cell removal **(Methods)**, yielded a final scRNA map of 279,155 cells across 14 datasets comprising 118 samples, 3 sequencing platforms (10X, Smart-seq2 and Cel-seq2) and 3 atherosclerosis-prone arterial beds **(Fig. 2a and Supplementary Fig. 1i)**.

**Figure 1.**
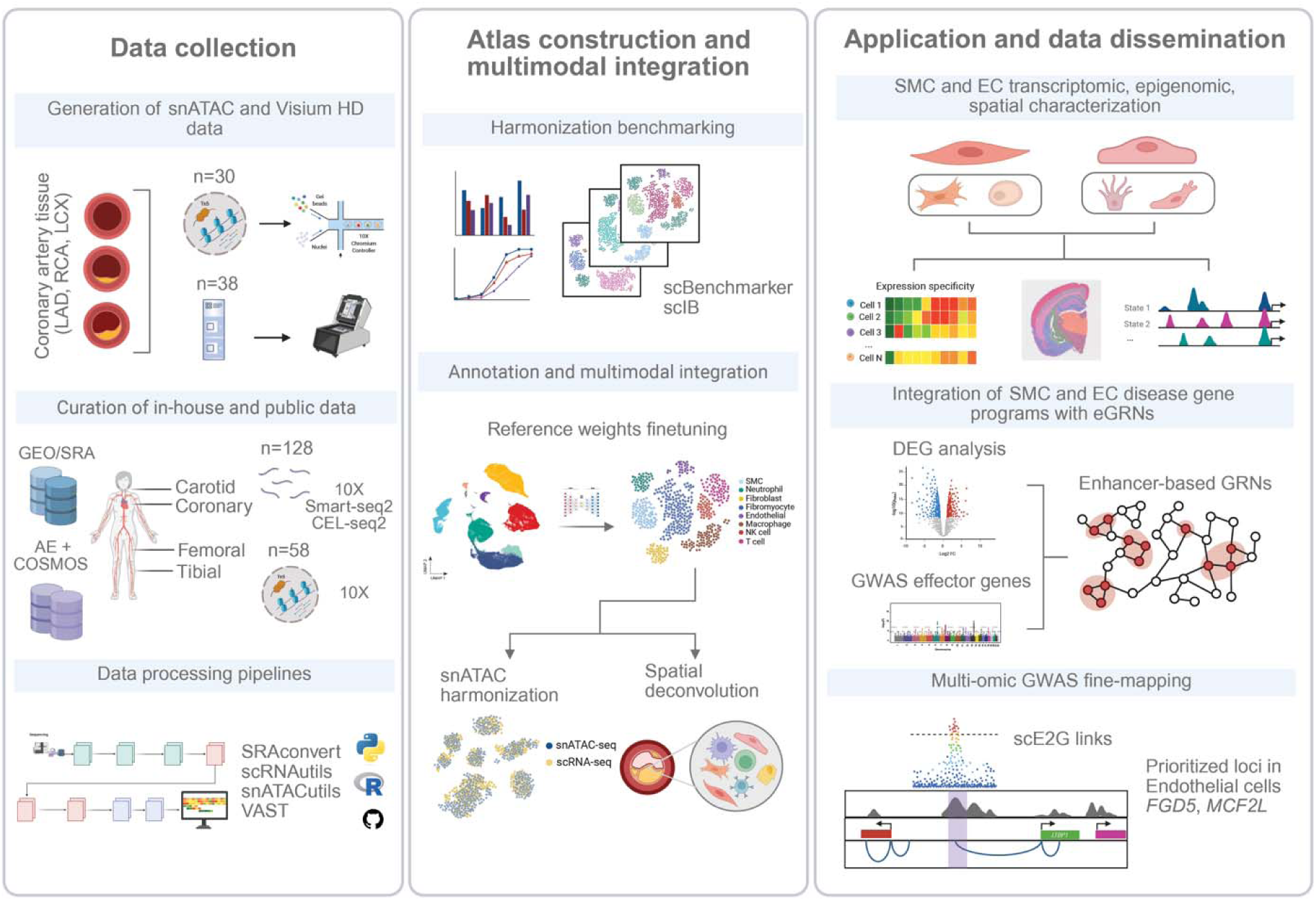
Overall workflow of the study. MetaPlaq includes published, in-house and newly generated datasets from single-cell and spatial assays across multiple atherosclerosis-relevant arterial beds. We implement pipelines for robust quality control at the sample and cell levels across included omic layers. Upon quality control, benchmarking of different integration algorithms was performed to build the atlas in a hierarchical manner and add detailed cell annotations. Presented applications included spatial validation of transcriptomics findings, construction of multimodal GRNs underlying disease-relevant transcriptional programs and multi-omics GWAS fine-mapping. LAD, left anterior descending artery; RCA, right coronary artery; LCX, left circumflex artery; AE, AtheroExpress; COSMOS, Coronary artery single-cell multi-omics study; GEO, Gene Expression Omnibus; SRA, Sequence Read Archive; 10x, 10x Genomics; SMC, smooth muscle cells; EC, endothelial cells; scIB, single-cell integration benchmarking; GRN, gene regulatory network; scE2G, single-cell enhancer-to-gene links.

**Figure 2.**
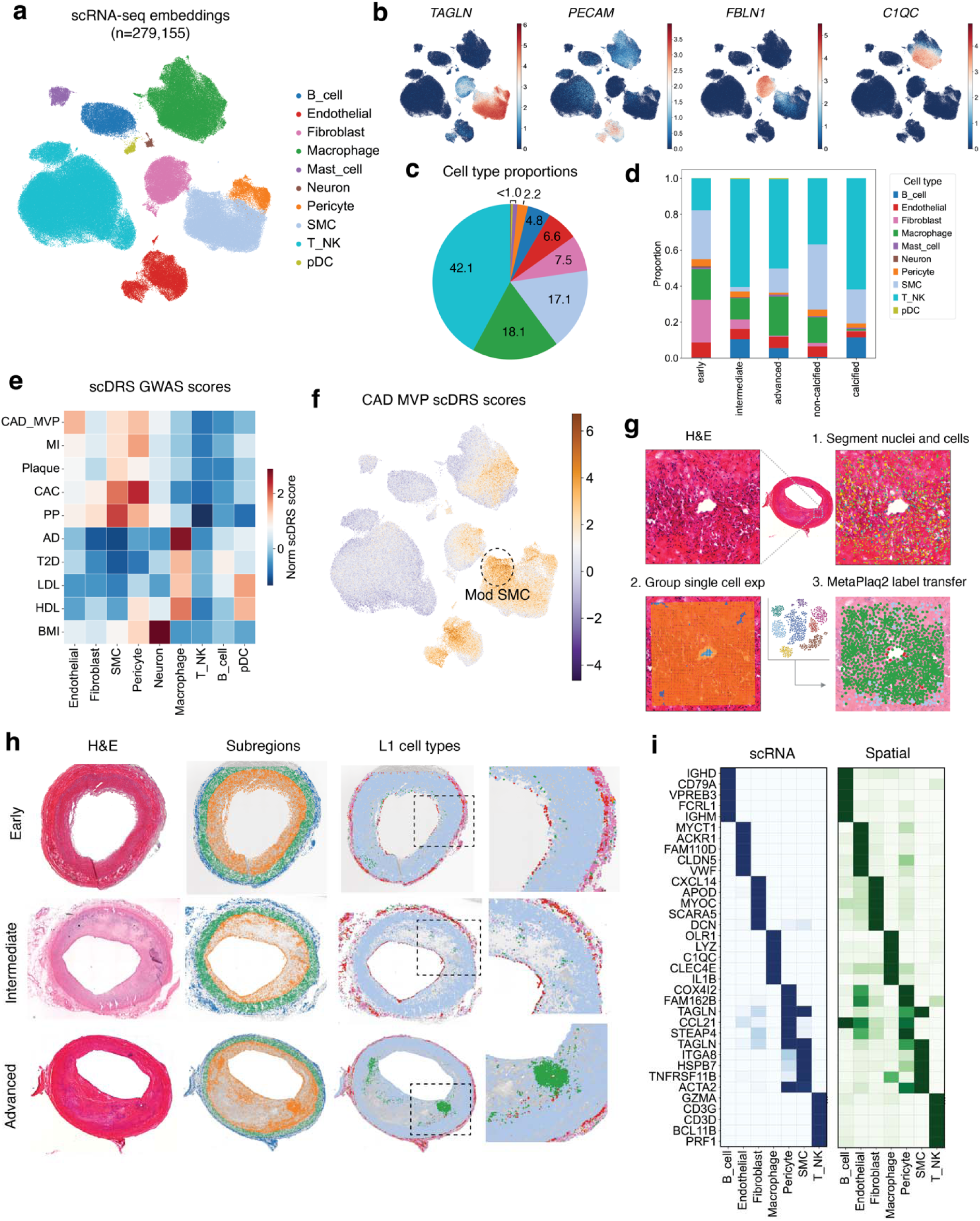
Integration of single cell and spatial transcriptomics maps major vascular and immune compartments in atherosclerosis. **a,** UMAP embeddings of 279,155 cells generated from scArches-based integration across 128 samples and 14 datasets, with cells colored based on their level 1 (L1) annotations. **b,** UMAP plot showing normalized expression of canonical vascular and immune marker genes *TAGLN* (SMCs), *PECAM1* (ECs), *FBLN1* (Fibroblasts), *C1QC* (Macrophages). **c,** Pie chart showing the percentage distribution of vascular and immune L1 annotations in **a**. **d,** Stacked barplot showing the distribution of L1 annotations across atherosclerotic lesion stage (early, intermediate, advanced, calcified, non-calcified). Lesion classifications were added based on author-provided labels or histology, when available. **e,** Heatmap displaying the mean normalized single cell disease relevance scores (scDRS) scores for cardiovascular and immune GWAS traits across L1 annotations. CAD: Coronary artery disease; MI: Myocardial infarction; CAC: Coronary artery calcification; PP = Pulse pressure; AD: Alzheimer’s disease; T2D: Type 2 diabetes; LDL and HDL: low/high density cholesterol; BMI: Body mass index. **f,** Representative UMAP plot showing per-cell scDRS normalized scores for CAD. Orange depicts cells enriched for gene-level association signals; whole non-relevant cells are colored in dark blue. Modulated SMCs, broadly identified by loss of canonical differentiation markers are circled. **g,** Schematic of pipeline for cell segmentation of Visium HD data and label transfer from scRNA reference. Inset indicates segmented nuclei within the plaque shoulder of an advanced coronary artery lesion, single-cell assigned expression and the scANVI cell type transferred from the RNA reference. **h,** Grid plot showing representative H&E images of coronary artery samples used for spatial transcriptomics analyses across early, intermediate and advanced lesion stages. The second column indicates anatomical regions (adventitia, media, intima). The third column depicts RNA L1 labels transferred onto Visium HD data shown in **g**. Insets highlight macrophage infiltration across the three lesion stages. **i,** Heatmaps comparing mean normalized expression values of scRNA L1 Wilcoxon markers to normalized expression of their counterparts in high-resolution spatial data within the corresponding cell class.

After QC and harmonization, we assigned level 1 (L1) annotations using canonical markers and automated label transfer, identifying 9 major cell compartments **(Fig. 2a,b)**. Manual labels revealed exceptional concordance with predictions from our published reference^9^, both globally and within individual cell types **(Supplementary Fig. 2a,b)**. The atlas was enriched for lymphoid origin, likely reflecting inclusion of CD45-enriched datasets, with SMCs as the next most abundant population **(Fig. 2c).** Advanced and calcified lesions showed increased immune cell content, particularly T/NK and B cells **(Fig. 2d)**. Transcription factor (TF) activity inference identified canonical regulators, including MYOCD and TCF21 in SMCs and ERG in ECs, as well as candidate regulators such as NFIL3 and TRPS1 **(Supplementary Fig. 2c)**. Gene set enrichment analyses (GSEA) of top marker genes supported the expected biological functions of each compartment **(Supplementary Fig. 2d and Supplementary Table 2)**. Integrating MAGMA^23^ gene-level associations with scDRS^24^ showed that vascular wall populations, particularly, SMCs, ECs and pericytes, were most strongly enriched for CAD, MI, and CAC, prioritizing these populations for downstream analyses **(Fig. 2e,f)**.

### High resolution spatial map of human coronary artery atherosclerosis

To define the anatomical context of L1 populations, we generated Visium HD spatial transcriptomics profiles from human coronary arteries with neointimal thickening (n=10), intermediate lesions (n=10) and advanced lesions (n=18) **(Supplementary Fig. 3a-c)**. We grouped multi-bin signals by underlying cells using bin2cell^25^ and annotated cell-level spatial profiles with SCANVI^26^ using our scRNA reference **(Fig. 2g, Methods)**. Subclinical lesions were composed mainly of fibroblasts, SMCs and ECs, with few macrophages in the neointima. In contrast, intermediate and advanced lesions showed a more complex mixture of vascular and immune cells: including SMC accumulation in the fibrous cap and macrophages, with fewer B cells, surrounding the necrotic core and plaque shoulders **(Fig. 2h)**. ECs localized both within plaques near immune infiltrates and in the adventitia, consistent with lymphatic endothelium or neovessel formation. SMCs showed the largest number of highly variable genes, L1 marker genes localized to expected compartments **(Supplementary Fig. 3d-f)** and spatial and single-cell profiles were highly concordant across lesion stages **(Fig. 2i)**, indicating that our spatial dataset captures cell-type heterogeneity during atherosclerosis progression.

### Immune and mural transcriptomic heterogeneity

We surveyed the major L1 immune and mural cell compartments using lineage-specific variable features **(Supplementary Fig. 4, Methods).** Manual annotations included markers of lymphoid, myeloid, and pericytes from the literature. A summary of the most representative cell subpopulations is provided in **Supplementary Note 1.**

### SMC and endothelial cell heterogeneity

Given the etiologic relevance of SMCs and ECs, we next resolved their transcriptional and spatial heterogeneity. Subclustering of SMCs identified 9 states, including contractile, fibromyocytes (FMCs), osteochondrogenic-like (fibrochondrocytes; FCs) and foam-like SMCs **(Fig. 3a)**. These states were validated by enrichment of lineage-traced murine SMC signatures from a recent trajectory dataset^10^ **(Fig. 3b)**, with particularly strong concordance between the murine CMC program and our manually defined FC state **(Fig. 3b and Supplementary Fig. 5a)** indicating cross-species conservation of osteochondrogenic programs. To compare modulated SMC states, we quantified transcriptional similarity by computing cosine similarities and centroid distances in PCA space. This showed that foam-like SMCs were transcriptionally closer to FCs than to contractile SMCs or FMCs **(Fig. 3c and Supplementary Fig. 5a,b),** potentially suggesting a common progenitor or transdifferentiation from one state to the other.

**Figure 3.**
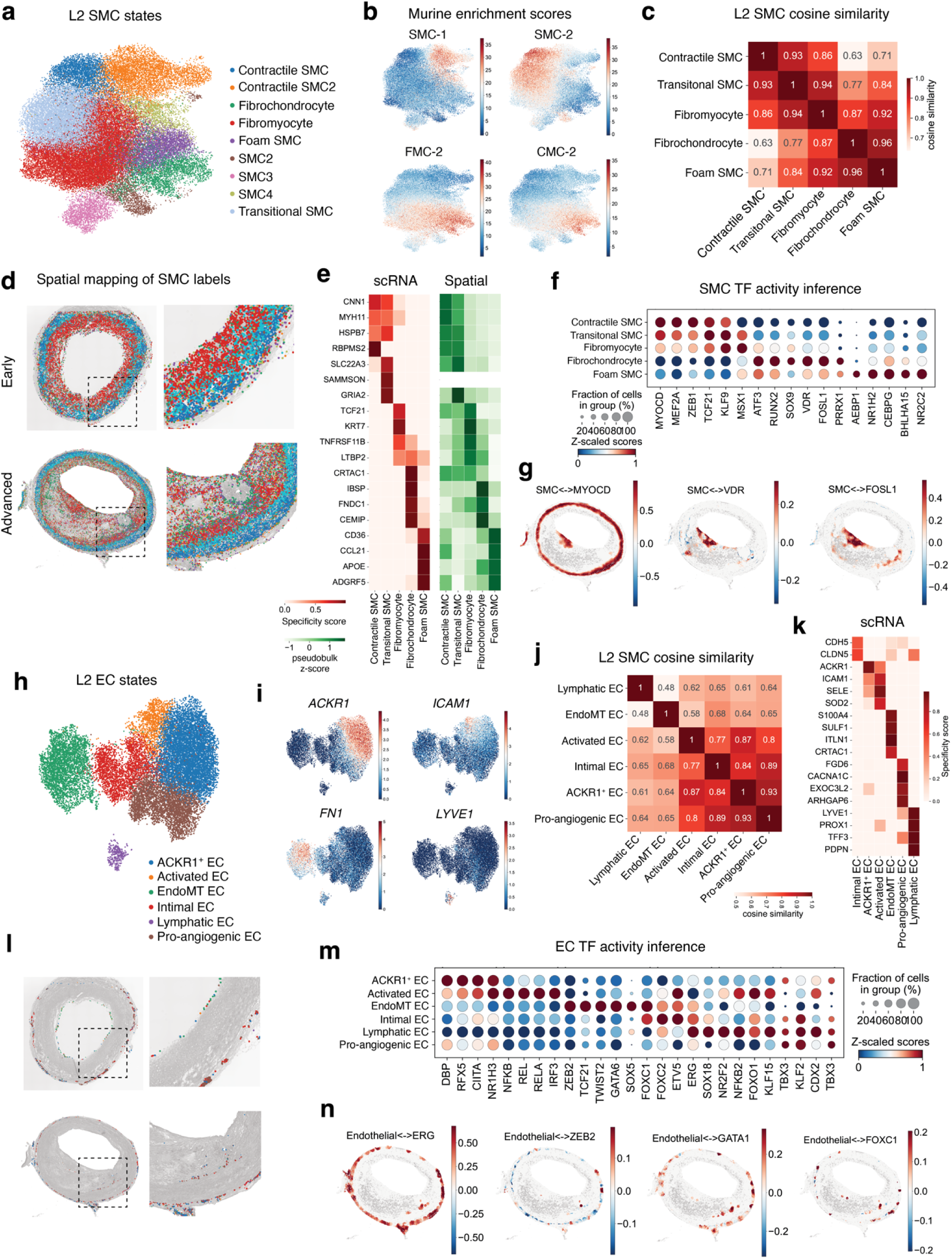
Single-cell and spatially resolved SMC and endothelial cell states. **a,** UMAP embeddings of SMC level 2 (L2) annotations encompassing 47,808 cells following the subclustering workflow described in **Methods**. **b,** UMAP of the SMC compartment showing enrichment of murine lineage-traced SMC signatures including chondromyocytes (CMC). Murine gene signatures were converted to human nomenclature and one-to-one orthologs were retained to build the final gene sets for enrichment with the decoupler package. **c,** Cosine similarities between the L2 SMC transcriptomic profiles. The mean normalized expression was computed across each gene and L2 annotation prior to similarity computation. **d,** Spatial mapping of L2 SMC annotations within early and advanced coronary lesions transferred from the scRNA reference onto Visium HD data. Granular labels show a gradient of phenotypic modulation from contractile SMCs in the media to FMCs through a transitional state, in addition FCs within the plaque. **e,** Heatmaps comparing expression specificity coefficients (ESμ) of scRNA SMC annotations to pseudobulked Z-scores of their counterparts in high-resolution spatial data within the corresponding cell class. Genes with low counts (below the third decile of expression) were removed to increase robustness of signal detection. **f,** Dot plot visualizing inferred transcription factor (TF) activity scores of SMC L2 annotations in the scRNA data using the univariate linear modeling framework from decoupler. Scores shown in the dot plot were standardized with Z-score scaling and dot size depicts fractions of cells exhibiting activity of the target TF. **g,** Spatial plots showing spatially weighted TF activity scores of factors in **g** restricted to SMCs**. h,** UMAP embeddings plot showing Endothelial (EC) L2 annotations encompassing 18,518 cells annotated as described in **Methods**. **i,** UMAP plot showing normalized expression of L2 EC markers *ACKR1* (ACKR1+ and Activated EC), *ICAM1* (Activated EC), *FN1* (EndoMT), *LYVE1* (Lymphatic EC). **j,** Cosine similarities measured across L2 EC scRNA profiles. Similarity coefficients were computed as described in **c.** **k,** Heatmap showing ESμ coefficients of highly specific EC L2 markers from the scRNA data computed as described in **e.** **l,** Spatial mapping of L2 EC annotations within early and advanced coronary lesions with labels transferred as described in **d**. **m,** TF activity scores of EC L2 annotations computed as described in **g**. **n,** Spatial plots showing spatially weighted regulon activity scores of factors shown in **n** restricted to EC.

We next mapped these SMC states to their anatomical locations using high resolution spatial transcriptomics. Training a scANVI model on the integrated MetaPlaq reference localized contractile SMCs to the medial layer, transitional states to the medial/intimal interface and FMCs primarily to the neointima. In advanced lesions, FCs and foam-like SMCs localized to vulnerable plaque regions, including plaque shoulders, whereas healthy and subclinical lesions contained few such states **(Fig. 3d and Supplementary Fig. 5c)**. To nominate state-specific programs we used the CELLEX framework^27^, which integrates 4 expression-specificity metrics into a robust specificity score (ESμ) **(Methods)**. This analysis identified canonical SMC genes such as *MYH11* and *MYOCD*, and highlighted candidate markers of FCs (*FNDC1, CEMIP, THY1 and FAP)* and foam-like SMCs (*APOE*, *APOC1*, *AGT*, *ADGRF5*, *DIO2* and *CD36)* **(Fig. 3e and Supplementary Table 3)**, with strong concordance to pseudobulked expression across L2 states **(Fig. 3e)**. Regulon analysis with decoupler^28^ confirmed high MYOCD and MEF2A activity in contractile SMCs, whereas FCs showed increased RUNX2, SOX9, FOSL1 and VDR activity. Intriguingly, foam-like SMCs were enriched for AEBP1 and NR1H2 activity, consistent with known lipid handling programs from adipocytes **(Fig. 3f and Supplementary Fig. 5d)** ^29^; ^30,31^.

These TF activities were constrained to the medial layer and plaque of advanced lesions **(Fig. 3g),** as depicted by spatially weighted cosine similarity of these TFs expression and SMCs location.

Subclustering of endothelial cells identified 6 states, including a cluster with strong expression of the atypical chemokine receptor-1 (*ACKR1*+), activated, pro-angiogenic, EndoMT and lymphatic ECs **(Fig. 3h)**. Activated ECs expressing *ICAM1* also expressed *ACKR1,* suggesting that *ACKR1*+ ECs may represent an early dysfunctional EC state **(Fig. 3i),** consistent with the known binding of this receptor to pro-inflammatory and angiogenic chemokines^32,33^.

Consistently, cosine similarity and euclidean distance calculations showed that *ACKR1*+ ECs were more similar to activated and pro-angiogenic ECs than to EndoMT and lymphatic ECs **(Fig. 3j and Supplementary Fig. 5f)**. ESμ analysis identified candidate EndoMT markers, including *CRTAC1*, *GATA6*, *SULF1 and ITLN1* **(Fig. 3k and Supplementary Table 4)**, whereas pro-angiogenic ECs were marked by genes such as *FGD6* and *EXOC3L2,* which have established roles in vascular development and disease-related angiogenesis^34^.

Spatial mapping showed that early intimal lesions contained a mixture of *ACKR1*+, activated and EndoMT ECs, whereas advanced lesions showed greater localized pro-angiogenic, *ACKR1+* and EndoMT cells to plaque shoulders **(Fig. 3l),** consistent with increased EC migration and proliferation during lesion progression. Intriguingly, transcriptomics-based regulon analysis identified NFκB activity only in activated ECs, consistent with reported activity of this TF in human aortic Endothelial cells (HAECs) stimulated with IL1-B^35^. Intriguingly, we also found striking enrichment of TCF21, SOX5, ZEB2 and FOXC1 regulon activity in EndoMT ECs **(Fig. 3m and Supplementary Fig. 5g,h),** with the last 2 localized to the adventitia and plaque regions of advanced coronary lesions **(Fig. 3n).** Altogether, layering scRNA and Visium HD data provides a spatial context for the activity of regulatory programs linked to SMC and EC phenotypic transitions, including osteogenic and mesenchymal states, respectively.

### Differential gene expression associated with atherosclerosis lesion stage

To define disease relevant gene signatures, we first analyzed Visium profiles across atherosclerosis lesion progression using DEseq2^36^. Comparison of advanced versus early lesions identified 199 DEGs, including 114 upregulated genes (**Fig. 4a and Supplementary Table 5**). Genes upregulated in advanced lesions, such as *C1QA, LIPA*, *CD163,* and *IGKC,* were consistent with increased immune responses during lesion progression **(Fig. 4a,b and Supplementary Fig. 6a-c).** To provide spatial context, we used PROGENy^37^ to infer single-cell pathway activity, which showed increased NFκB, JAK-STAT and TNF signaling in advanced lesions, with strongest activity at plaque shoulders and surrounding the necrotic core (**Fig. 4c,d**). Together, these findings support established roles for infiltrating myeloid cells and inflammatory signaling in plaque progression.

**Figure 4.**
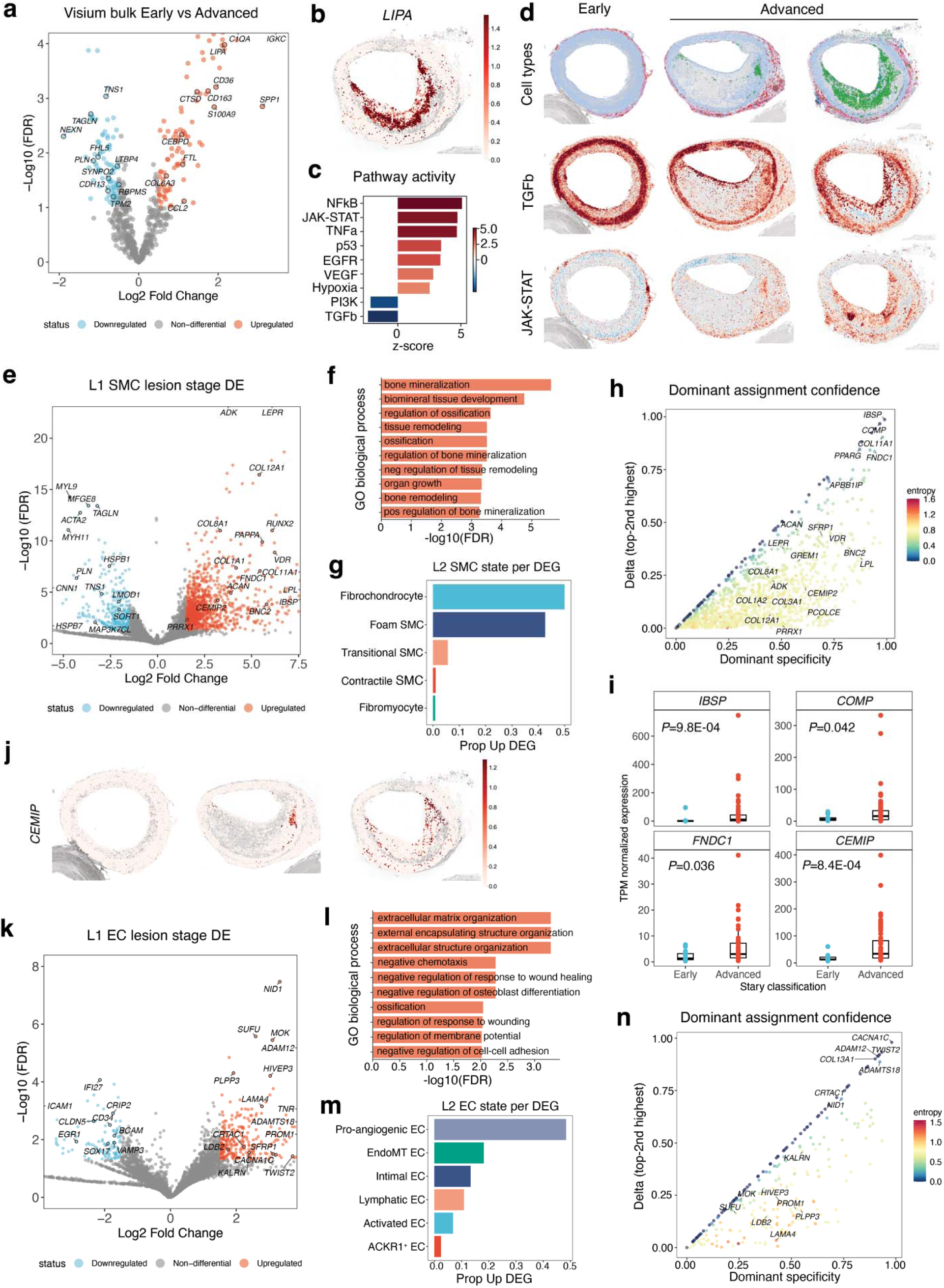
Disease-associated gene signatures by atherosclerotic lesion stage. **a,** Volcano plot showing pyDeseq2 analyses in Visium HD data comparing advanced to early stage coronary atherosclerotic lesions. Differentially expressed genes (DEGs) were identified under the thresholds (FDR < 0.1 and |log_2_ (FC)| > 1) and are depicted as light red (upregulated) and blue (downregulated). **b,** Spatial plot mapping normalized expression of lysosomal acid lipase (*LIPA)*, a representative upregulated gene in coronary advanced lesion samples. **c,** Bar plot showing enrichment of Progeny pathways within pseudobulked advanced vs early coronary lesions. **d,** Spatial plots showing the single-cell resolution enrichment/localization of representative pathways in **c** across coronary samples representative of early and advanced lesions. **e,** Volcano plot of L1 SMC DEGs in advanced compared to early atherosclerotic lesions from scRNA data. DEGs were identified under the thresholds (FDR < 0.05 and |log_2_ (FC)| > 1.5) and are colored as light red (upregulated) and blue (downregulated). **f,** Bar plot showing gene ontology biological processes (GO BP) enriched within the top 100 SMC upregulated genes in advanced lesions. Genes were ranked by log_2_ (FC) and enrichment was conducted using gProfiler2 with the hypergeometric test. Top GO BP terms are listed in the *y* axis and FDR values shown in the *x* axis are adjusted for multiple testing. **g,** Bar plot showing the proportion of upregulated genes in **e** assigned to each SMC L2 annotation. Assignments were done based on the highest ESμ score across considered L2 states. **h,** Scatter plot showing the degree of DEG sharing across L2 annotations using information theory. Each dot represents a gene. The *x* axis shows the ESμ score while the *y* axis shows the computed delta Δ (difference between the 2 highest scores). Genes are colored by their corresponding Shannon entropy of specificity across L2 states. **i,** Bulk RNA-seq expression of *IBSP*, COMP, *FNDC1* and *CEMIP* in coronary arteries from early (n = 13) and advanced lesion samples (n = 59). The *y* axis represents normalized expression counts (TPMs). *P* values were calculated using a non-parametric Wilcoxon rank-sum Test. Boxplots represent the median and the inter-quartile (IQR) range with upper (75%) and lower (25%) quartiles shown, and each dot represents a separate individual. **j,** Spatial plot showing normalized expression of *CEMIP* in early, intermediate and advanced coronary lesions. **k,** Volcano plot of L1 EC DEGs genes in advanced vs early atherosclerotic lesions from scRNA data. Significance thresholds and color schemes are similar to those in **e**. **l,** GO BP ontology terms of the top 100 EC genes upregulated in advanced lesions. Ranking and enrichment were conducted as described in **f.** **m,** Bar plot showing the proportion of upregulated genes in **k** assigned to each L2 EC annotation as done in **g**. **n,** Scatter plot showing the degree of DEG specificity across EC L2 annotations using information theory as described in **h**.

Given that genes downregulated in advanced lesions were linked to contractility and ECM-related terms **(Supplementary Fig. 6c)**, we next investigated disease-associated transcriptional programs by pseudobulking SMCs and ECs **(Supplementary Fig. 7a-c and Methods)**. We identified 1,503 DEGs, of which 1,178 were upregulated in advanced lesions **(Fig. 4e and Supplementary Table 6).** Upregulated genes included *IBSP*, *RUNX2*, *FNDC1*, *COMP,* and *ADK*—a coronary artery calcification (CAC) GWAS gene^38^—and were enriched for osteochondrogenic processes such as bone mineralization and ossification **(Fig. 4f and Supplementary Table 7)**, pointing to SMC-driven calcification in advanced lesions.

Next, we used information theory to formally distinguish whether differential signatures were driven by a specific L2 state or shared across annotations. First, we assigned each DEG to the L2 state with the largest ESμ value, which revealed a strong contribution of FCs and Foam-like SMCs among upregulated genes **(Fig. 4g)** while downregulated genes were biased towards contractile SMCs and FMCs **(Supplementary Fig. 7d,e).** A substantial fraction of upregulated DEGs were shared across two or more L2 SMC states (e.g., *FN1, LUM*) rather than restricted to a single state, reflecting general phenotypic modulation^9^. To formally distinguish shared and specific L2 markers, we computed Shannon Entropy values using L2 ESμ coefficients across each DEG **(Methods),** which revealed genes like *IBSP, PPARG* and *FNDC1* **(Fig. 4h)** unique to FCs and foam-like SMCs. Increased entropy in genes like *COL8A1* points to common gene programs between two or more states, potentially due to programs activated during earlier stages of phenotypic modulation. Intriguingly, upregulated DEGs with low-medium entropy in SMCs showed increased expression in coronary artery bulk RNA-seq samples **(Fig. 4i)**, in addition to marked localization to plaque regions of advanced lesions, as shown by *CEMIP* in our Visium data **(Fig. 4j)**.

We employed a similar approach to ECs and identified 401 DEGs, of which 289 were upregulated in advanced lesions. Genes increased in advanced lesions included the candidate EndoMT marker *CRTAC1* and *PLPP3* **(Fig. 4k and Supplementary Table 8)**, a CAD GWAS gene linked to altered vascular barrier integrity^39^. These upregulated genes were enriched for extracellular matrix remodeling and heart development terms **(Fig. 4l and Supplementary Table 9)**. In contrast, downregulated DEGs included canonical EC markers such as *CLDN5* and *EGR1*, and were associated with leukocyte infiltration signatures **(Supplementary Fig. 7f,g)**, pointing to endothelial dysfunction as an early hallmark of atherosclerosis. Upregulated EC DEGs mapped to pro-angiogenic and EndoMT states and were upregulated in advanced coronary lesions **(Figure 4m and Supplementary Fig. 8a,b).** Furthermore, they showed intermediate entropy values, indicating substantial sharing across EC phenotypic states **(Figure 4n)**.

### High-resolution chromatin accessibility atlas of human atherosclerosis

To define *cis*-regulatory programs in athero-relevant cell types, we analyzed snATAC data from 82,396 nuclei, harmonizing 44,324 nuclei from public datasets with 38,072 nuclei from newly generated HCA libraries **(Supplementary Fig. 9a-c and Supplemental Table 10)**. Across 4 arterial beds, GLUE^40^ integration outperformed spectral embedding, Harmony^20^, and PeakVI^41^, and produced the best cell type alignment across RNA and ATAC modalities while preserving biological structure **(Fig. 5a, Supplementary Fig. 9d-g)**. Transfer of transcriptomics labels, refined by inferred gene activity, identified 8 major vascular and immune populations **(Fig. 5b-d).** Gene expression profiles were highly correlated with inferred gene activity across major cell types. **(Supplementary Fig. 9j-l).** We identified 515,498 open chromatin regions (OCRs) in this atlas, the largest chromatin map reported in athero-relevant tissues (**Methods**). Motif enrichment analysis of L1 cell type specific OCRs uncovered canonical regulators such as SRF and MEF2 in SMCs, SOX factors in endothelial cells; SPI factors in macrophages and T-Box, RUNX and EOMES factors in T/NK cells **(Fig. 5e)**. Importantly, motif deviations correlated with local accessibility of ESμ-derived cell type features (e.g., HSPB7 and MYCT1 in SMC and ECs, respectively) **(Fig. 5f-h)**.

**Figure 5.**
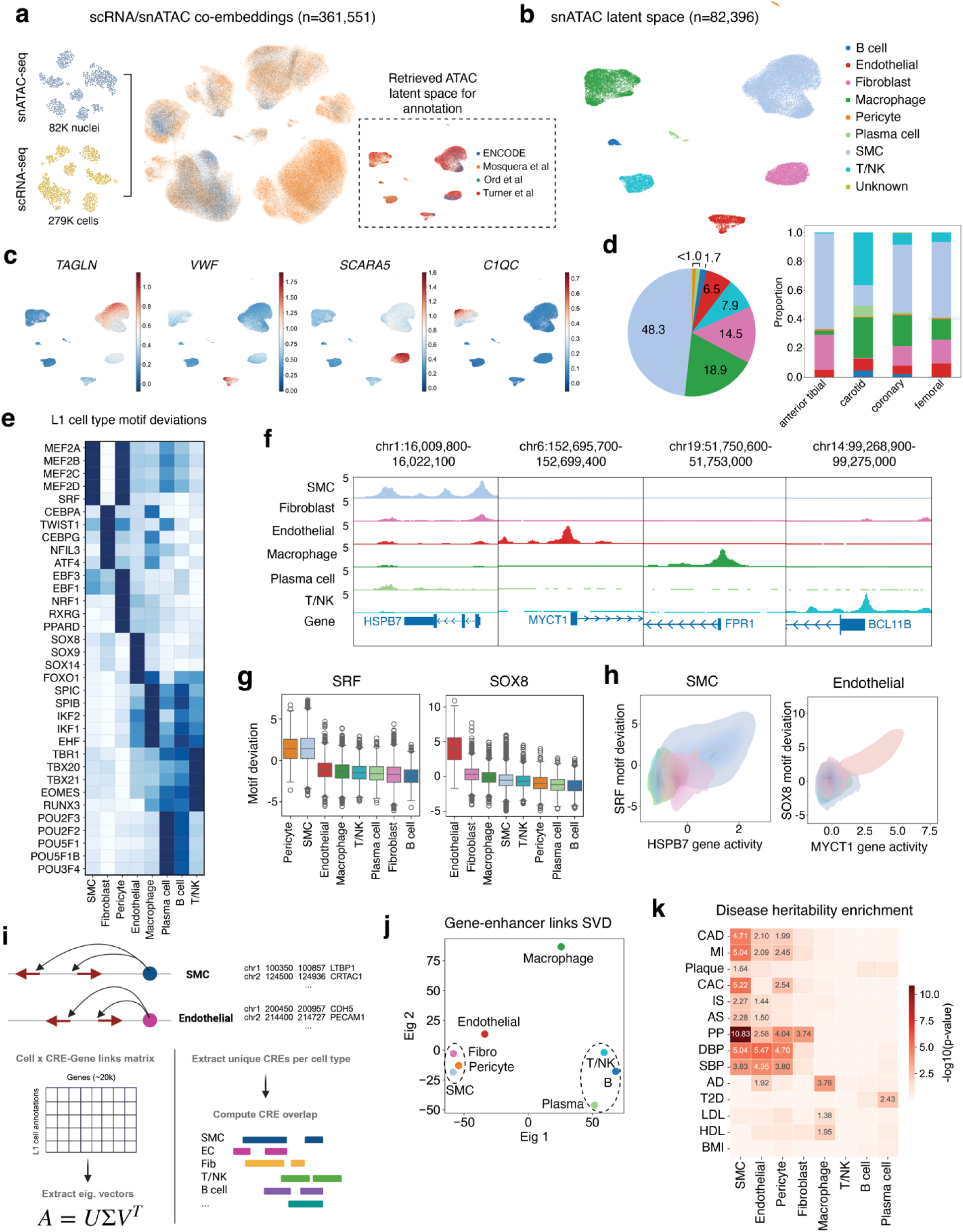
High resolution chromatin accessibility atlas of human atherosclerosis. **a,** Schematic showing multimodal harmonization of 361,551 cells across 88 samples. scRNA-seq and snATAC-seq modalities were co-embedded using GLUE in supervised mode. Dashed box depicts post-integration ATAC latent space colored by dataset and used for annotations. **b,** UMAP embeddings showing level 1 (L1) annotations for 82,396 nuclei in snATAC-seq data. **c,** UMAP plots of snATAC latent space showing normalized gene activity scores of canonical vascular and immune markers *TAGLN* (SMCs), *PECAM1* (ECs), *FBLN1* (Fibroblasts), *C1QC* (Macrophages). **d,** Pie chart showing the percentage distribution of L1 snATAC annotations (left). Stacked barplot showing the distribution of L1 annotations across arterial beds in the meta-analyzed snATAC reference (right). **e,** Heatmap showing mean motif deviation scores of top 7 marker transcription factors (TFs). Motif deviations were computed with pychromvar and marker TFs were obtained through a Wilcoxon rank-sum test. **f,** Genome browser tracks showing normalized accessibility at the transcription start site (TSS) of highly specific genes across L1 annotations, including *HSPB7* (SMC), *MYCT1* (EC), *FPR1* (Macrophage), *BCL11B* (T/NK). Specificity was defined based on ESμ coefficients computed across L1 scRNA populations. **g,** Motif deviation scores of TF markers SRF(SMC) and SOX8(EC) across L1 annotations. Boxplots represent the median and the inter-quartile (IQR) range with upper (75%) and lower (25%) quartiles shown. **h,** Density plots showing correlation of SMC and EC marker motif deviations in **g** to gene accessibility of highly specific SMC and EC genes, including *HSPB7* and *MYCT1*. **i,** Schematic showing identification of L1 cell-specific scE2G enhancers and downstream dimensionality reduction and overlap analyses. **j,** Scatter plot showing singular value decomposition (SVD) factorization of a matrix where each entry represents the number enhancer-gene links per L1 annotation. Eig 1 represents eigenvector 1 which explains most of the variance in enhancer connections. **k,** Stratified LD score regression (S-LDSC) applied to L1 differentially accessible regions (DARs) across cardiovascular and non-cardiovascular GWAS traits. Heatmap entries with numeric annotations depict cells passing the significance threshold for a given trait (*P* < 0.05 at - log_10_(*P*) = 1.301). CAD: Coronary artery disease; MI: Myocardial infarction; CAC: Coronary artery calcification; IS: Any ischemic stroke; AS: Any stroke; PP: Pulse pressure; DBP: Diastolic blood pressure; SBP: Systolic blood pressure; AD: Alzheimer’s disease; T2D: Type 2 diabetes; LDL and HDL: low/high density cholesterol; BMI: Body mass index.

Next, we applied scE2G^42^ to infer cell type-specific enhancer-gene interactions. Although less abundant than SMCs, endothelial cells and macrophages showed comparable or greater numbers of predicted links **(Supplementary Fig. 10a)**, implicating signal strength in addition to cell abundance. Singular value decomposition (SVD) of an enhancer-gene matrix and genomic overlap analyses showed greater enhancer sharing among SMCs, pericytes and fibroblasts than with endothelial cells, whereas lymphoid cells segregated from macrophages, consistent with a more divergent myeloid enhancer landscape **(Fig. 5i, j and Supplementary Fig. 10b)**. Differential accessibility analysis identified 187,771 differentially accessible regions (DARs) enriched for expected cell type functional ontology terms and localized predominately in intergenic and intronic regions 10-100 kb from the nearest TSS, consistent with known enhancer architecture **(Supplementary Fig. 11a-f)**. Immune DARs overlapped strongly with PBMC ENCODE profiles, whereas vascular DARs showed weaker enrichment **(Supplementary Fig. 11g)**, highlighting the lower representation of athero-relevant vascular states in public datasets. Finally, stratified LD score regression analysis (S-LDSC)^43^ with multiple vascular and immune GWAS traits **(Supplementary Table 11)** showed that SMC DARs were most enriched for CAD^7^, pulse pressure (PP)^7,44^, CAC^38^ and myocardial infarction (MI)^45^ signals, followed by a significant contribution from EC and pericyte DARs (**Fig. 5k**), supporting key etiologic roles reported in **Fig. 2e**.

### Chromatin accessibility landscape of SMC and endothelial cells

Given the heritability enrichment of SMC and endothelial cell DARs for cardiometabolic traits, we next resolved chromatin states for these cell types. Within ECs, we resolved 3 states: *ACKR1*^+^/*ICAM1^+^* immune and pro-angiogenic ECs, EndoMT ECs and Lymphatic ECs **(Fig. 6a).** Canonical EndoMT markers including *ZEB2*, along with ESμ nominated markers such as *CRTAC1* and *SULF1*, showed high accessibility in the EndoMT cluster **(Fig. 6a,b and Supplementary Fig. 12a,b)**. The *ACKR1+* cluster was enriched for ETS factors (e.g., ETV1/2, ELF1) in addition to immune related TFs such as IKZF2, previously linked to pathologic angiogenic responses^46^. By contrast, EndoMT OCRs were enriched for GATA6, FOXC1, and TRPS1 **(Fig. 6b,c)**, all established regulators of SMC differentiation and phenotypic switching^47–49^. Their activity in this population is consistent with the acquisition of mesenchymal features characteristic of other lineages and suggests they may also drive endothelial-to-mesenchymal transition in human atherosclerosis.

**Figure 6.**
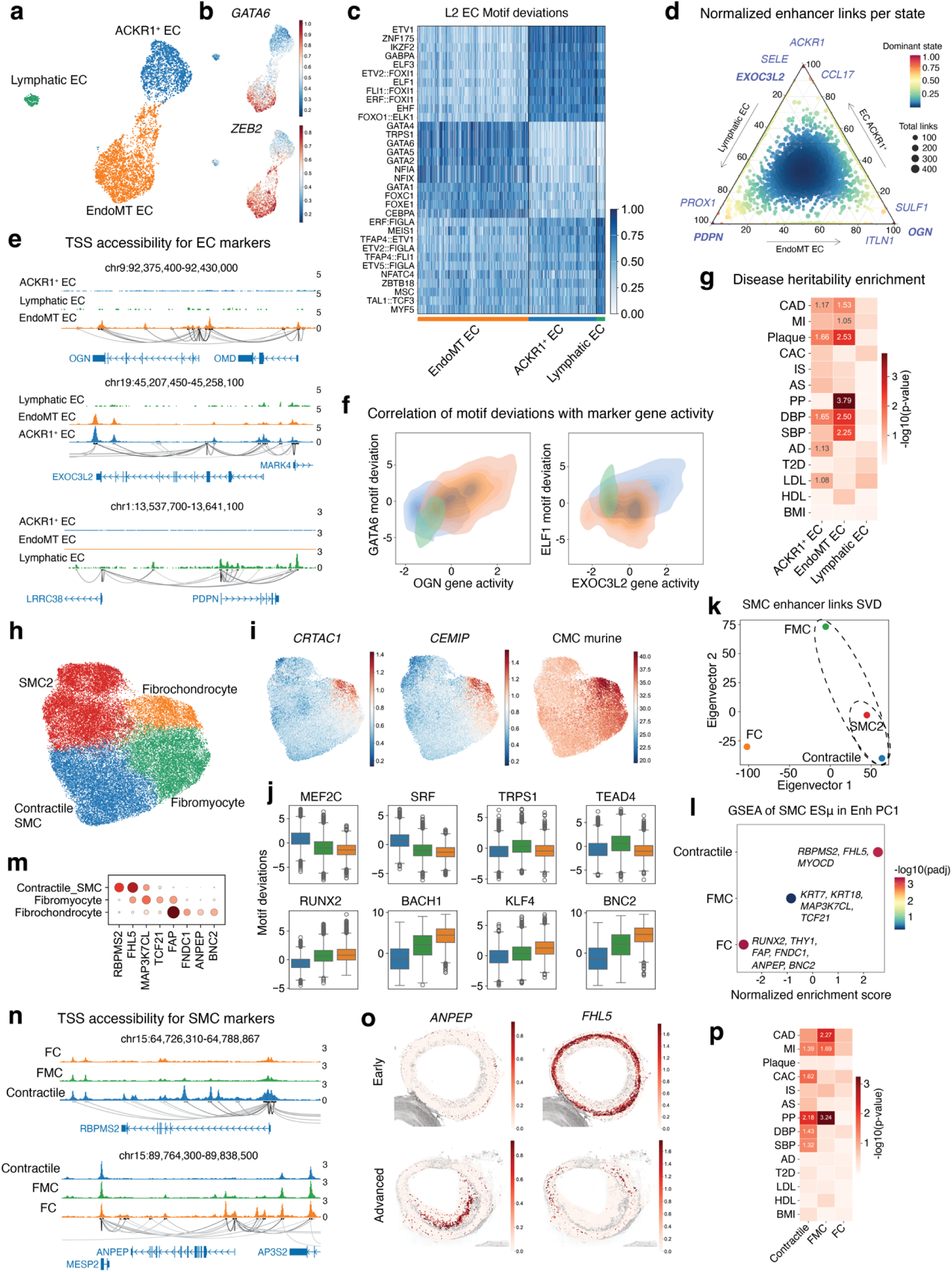
Chromatin atlas reveals cis-regulatory landscape of SMC and EC states. **a,** UMAP embeddings of EC level 2 (L2) snATAC annotations encompassing 5,388 nuclei. Nuclei were annotated using the subclustering workflow described in **Methods**. **b,** UMAP plots showing normalized gene activity scores of Endothelial-to-mesenchymal (EndoMT) markers *ZEB2* and *GATA6*. Activity scores were computed by aggregating the accessibility Tn5 insertion counts within a given gene body using snapatac2, and subsequently log-normalized with scanpy. **c,** Heatmap showing mean motif deviation scores of top 10 marker TFs across L2 EC clusters. Motif deviations were calculated with pychromvar. Marker TFs were obtained by performing a Wilcoxon Rank Sum test with scanpy. **d,** Ternary plot showing row-normalized number of enhancer connections across the entire EC transcriptome. Dots depict individual genes and are colored by their specificity value across EC L2 annotations. Dot size reflects the number of enhancer connections targeting a given gene. **e,** Genome browser tracks showing normalized accessibility of genes with differential enhancer loads across EC L2 annotations shown in **d**. **f,** Density plots correlating deviations of top EC L2 motifs from **c** to gene accessibility profiles of top specific markers identified in **d.** **g,** Stratified LD score regression (S-LDSC) applied to L2 EC differentially accessible regions (DARs) across cardiovascular and non-cardiovascular GWAS traits. Heatmap entries with numeric annotations depict cells passing the significance threshold for a given trait (*P* < 0.1 at - log_10_(*P*) = 1). CAD: Coronary artery disease; MI: Myocardial infarction; CAC: Coronary artery calcification; IS: Any ischemic stroke; AS: Any stroke; PP: Pulse pressure; DBP: Diastolic blood pressure; SBP: Systolic blood pressure; AD: Alzheimer’s disease; T2D: Type 2 diabetes; LDL and HDL: low/high density cholesterol; BMI: Body mass index. **h,** UMAP embeddings of SMC L2 snATAC annotations encompassing 39,771 nuclei. Nuclei were annotated using the subclustering workflow described in **Methods**. **i,** UMAP plots showing normalized gene activity scores of FCs (*CRTAC1* and *CEMIP*) in the SMC subset from **h**. Right-most UMAP plot shows lineage-traced SMC enrichment scores for murine chrondromyocytes (CMCs). **j,** Motif deviation scores for top L2 SMC marker motifs from Wilcoxon Rank Sum tests. Boxplots represent the median and the inter-quartile (IQR) range with upper (75%) and lower (25%) quartiles shown. **k,** Scatter plot showing singular value decomposition (SVD) factorization of matrix counting enhancer links per SMC L2 annotation. Eig 1 represents the first eigen vector explaining most of the variance in enhancer connections. **l,** Dot plot showing gene set enrichment (GSEA) of SMC L2 gene sets in the 95th percentile of expression specificity (based on ESμ scores) within the first eigenvector loadings from **k**. GSEA was performed with the fgsea R package. The *x* axis depicts the normalized enrichment score (NES) computed based on a modified Kolmogorov-Smirnov (K-S) test. The *y* axis shows the SMC L2 input gene sets. Dots are colored by the -Log_10_(*P* value), computed by comparing the observed ES against a distribution of ES values obtained from random permutations of gene ranks. **m,** Dot plot visualizing normalized mean expression of representative L2 SMC-specific genes showing enrichment within the eigenvector 1 loadings from **j**. **n,** Genome browser tracks of SMC L2 states showing normalized accessibility of RBPMS2 (Contractile SMC) and ANPEP (FC) highlighted in **l** and **m**. **o,** Spatial plot showing normalized expression of *FHL5* and *ANPEP* in early and advanced coronary lesions. **p,** S-LDSC applied to SMC L2 DARs across cardiovascular and non-cardiovascular GWAS traits. Heatmap entries with numeric annotations depict cells passing the significance threshold for a given trait (*P* < 0.1 at -log_10_(*P*) = 1). Abbreviations for traits are described in **g**.

Interestingly, E2G predictions showed greater enhancer sharing between *ACKR1+* and EndoMT clusters than with the lymphatic cluster **(Supplementary Fig. 12c,d),** consistent with the notion that specific epigenetic programs drive arteriovenous EC specification^50^. Although most genes showed similar enhancer burdens across states, each population had modest differential enhancer-gene interactions pointing to activation of state-specific gene programs **(Fig. 6d)**.

Genes highlighted by ESμ analyses included *SULF1* and *OGN* in EndoMT ECs, *EXOC3L2* in *ACKR1*+ ECs and *PDPN* in lymphatic ECs; each showed marked differences in TSS accessibility, and gene body accessibility correlated with motif deviations for corresponding regulators **(Fig. 6d-f)**. We identified a total of 5,684 L2 EC DARs, which were enriched for processes including blood vessel morphogenesis in *ACKR1*+ ECs, and ECM organization and TGFB signaling in EndoMT ECs **(Supplementary Fig. 12e)**. S-LDSC analyses using these DARs showed that EndoMT captured a substantial fraction of the EC contribution to cardiometabolic trait heritability, including pulse pressure (PP), further supporting the mesenchymal features of this state **(Fig. 6g)**.

Among 39,771 SMCs, we identified 4 L2 states, including contractile SMCs, FMCs and FCs, using canonical and ESμ-derived markers **(Fig. 6h,i and Supplementary Fig. 13a,b).**

Annotations were cross-referenced against murine SMC signatures^10^, and our FC label showed strong concordance with nuclei with the highest murine CMC scores **(Fig. 6i and Supplementary Fig. 13c)**. Contractile SMC OCRs were enriched for SRF and MEF2 motifs, whereas we observed progressively greater enrichment of RUNX2, KLF4, BACH1/2 and BNC2 motifs along the SMC dedifferentiation axis **(Fig. 6j)**. Notably, BNC2 was among our SMC DEGs in **Fig. 4e** and was nominated in our multi-trait GWAS of CAD and subclinical atherosclerosis traits ^51^. E2G predictions showed slightly more CREs in contractile SMCs and FMCs than in FCs, and greater enhancer sharing between contractile SMCs and FMCs compared to FCs **(Supplementary Fig. 13d,e)**.

Given the strong overlap across L2 SMC accessibility profiles, we applied SVD to enhancer-gene link profiles, which separated SMCs and FMCs from FCs along the first eigenvector **(Fig. 6k)**. GSEA of ESμ-nominated L2 signatures showed enrichment of contractile markers such as *MYOCD, FHL5* and *RBPMS2* among features with positive loadings, whereas FC genes such as *FAP* and *ANPEP* were enriched among features with the highest negative loadings **(Fig. 6l and Supplementary Fig. 13f)**. These genes showed progressively increased TSS accessibility with FC modulation and were highly expressed in the neointima of advanced lesions compared to subclinical controls **(Fig. 6m-o).** SMC L2 DARs were enriched for terms such as regulation of muscle contraction, ECM organization and chondrocyte differentiation, and for CArG and RUNX1/2 motifs in contractile SMCs and FCs, respectively **(Supplementary Fig. 13g-i)**.

Furthermore, FMC DARs also showed the strongest enrichment for CAD and MI heritability **(Fig. 6p),** suggesting that osteogenic SMC programs reflect downstream responses rather than primary mediators of CAD genetic risk.

### Enhancer-driven disease-associated gene regulatory networks

To link disease-relevant SMC and EC programs to *cis*-regulatory landscapes, we prioritized cell-specific CAD GWAS loci by ranking genes by ESμ and intersecting them with CAD MAGMA gene sets **(Methods).** We define genes meeting both criteria—high cell-specific ESμ and CAD MAGMA significance—as cell-specific GWAS effector genes. This recovered known SMC and EC loci, including *PRDM16*^8,52^*, FHL5*^53^ and *FLT1*^34^ (**Fig. 7a**). In ECs, this set recovered 17% of the endothelial variant-to-gene-to-program (V2G2P) genes mapped by prior EC perturb-seq studies^34^, including loci like *CALCRL* and *FGD5* (**Supplementary Fig. 14 and Supplementary Table 12**). We also identified loci with less defined causal mechanisms and linked *CRTAC1* to EndoMT ECs and *PRDM16* to transitional SMCs (**Fig. 7a**).

**Figure 7:**
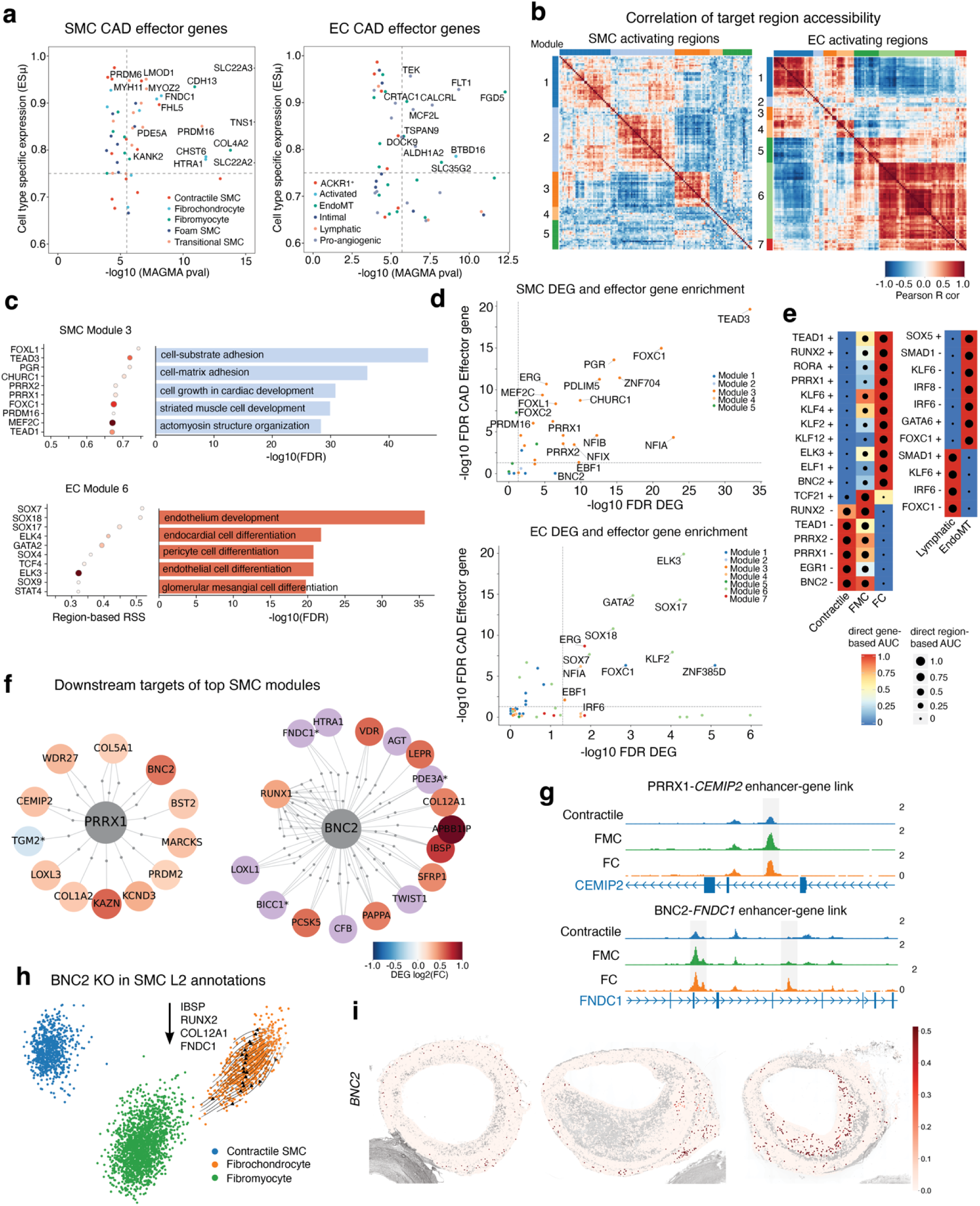
Enhancer-driven disease-associated gene regulatory networks. **a,** Scatter plots showing cell-specific GWAS effector genes. Effectors were obtained by intersecting a MAGMA-derived CAD gene set (top 1000 genes ranked by *p* value) with features in the 95th percentile of expression specificity (ESμ) for SMCs (left) and Endothelial cells (right). The *x* axis shows the -log_10_(MAGMA *P* values). Genes passing a significance threshold of ESμ = 0.75 and -log_10_(*P*)=5.3. **b,** Heatmap showing identified modules of predicted SMC- and EC-specific activating regulons (+/+). Module grouping was done using hierarchical clustering of pairwise L1 SMC (left) and EC (right) region-based AUC correlation values. **c,** Top 10 cell-specific L1 activating regulons (left) within SMC module 3 (top) and EC module 6 (bottom) ranked by region-based regulon specificity score (RSS). Dot size indicates number of genes (larger means regulon contains more genes) and dot color indicates number of regions (darker means regulon contains more regions) within individual regulons. Corresponding gene region enrichment analysis (GREAT) was performed on the union of regions among the top ten regulons shown (right). **d,** Enrichment of SMC-specific (top) or EC-specific (bottom) advanced lesion DEGs and effector genes in L1 activating regulons. Enrichment was done using a hypergeometric test. Dashed lines indicate the applied significance threshold (FDR < 0.05). Dot color indicates the SMC or EC module the regulon belongs to as identified in **a**. **e,** SCENIC+-predicted regulon activity within SMC (left) and EC (right) L2 annotations for regulating (+) and repressing (-) regulons. Box shade indicates enrichment of gene expression of target genes (GEX AUCell) and dot size indicates target region accessibility (AUCell). **f,** SCENIC+ identified networks for PRRX1 and BNC2 activating regulons in SMC L2 annotations. Transcription factors shown as the central grey node. Target regions represented as small grey dots and target genes shown as labeled, colored dots. Upregulated and downregulated DEGs in advanced lesions shown on blue/red color scale, SMC L1 effector genes shown in purple and those that are both DEGs and GWAS effector genes are marked with an asterisk. **g,** Genome browser tracks shown for target regions (putative enhancers) in TF-gene links with colors of tracks indicating SMC L2 states. PRRX1-*CEMIP2* target region (top) and BNC2-*FNDC1* target regions (bottom) are highlighted by grey boxes. Bottom track shows GENCODE v49 gene tracks of target gene. **h,** SCENIC+-predicted *in-silico* knockout of BNC2 within SMC L2 annotations. Cells are projected in PCA space derived from eRegulon gene AUC scores computed at the SMC L2 level and colored by the respective annotation. Direction and length of arrows indicate predicted trajectory and magnitude of shifts, respectively, in regulon activity following a BNC2 knockout, propagated across the gene regulatory network. Downward arrow signifies predicted downregulation of *IBSP*, *RUNX2*, *COL12A1*, and *FNDC1* following the BNC2 knockout. **i,** Spatial plot showing normalized expression of *BNC2* in early and advanced coronary lesions.

We next used SCENIC+^54^ to infer L1 enhancer-driven gene regulatory networks (eGRNs) **(Methods)**, revealing canonical regulators including ERG in ECs and MEF factors in SMCs (**Supplementary Fig. 14 and Supplementary Table 13**). Clustering highly active regulons by pairwise correlations using region-based AUC identified 5 SMC and 7 EC modules (**Fig. 7b, Supplementary Table 13**). While several modules were enriched for housekeeping processes, others captured cell identity and specialized functions; notably SMC module 3 and EC module 6 were enriched for pathways linked to SMC remodeling and endothelial dysfunction, respectively (**Fig. 7c**).

Given the limited overlap between DEGs and effector genes **(Supplementary Fig. 14)**, we tested enrichment of each gene set among eGRN targets individually to identify upstream regulators of these orthogonal disease programs. In SMCs, TEAD3 regulons were enriched for both DEGs and effector genes, whereas BNC2 and FOXC2 regulons were enriched for DEGs or effector genes, respectively (**Fig. 7d**). In ECs, FOXC1 and SOX17 were enriched for both gene sets, whereas IRF6^55^ was enriched only for DEGs (**Fig. 7d**). Several disease-enriched regulons also showed cell state specific activity; BNC2 had the highest accessibility in FCs, consistent with its observed motif deviation (**Fig. 6k and Fig. 7e**), alongside expected activity of RUNX2 in FCs and TCF21 in FMCs (**Fig. 7e**).

Inspection of SMC regulon targets identified PRRX1 regulation of *CEMIP2*^56^ through an intronic region with increased accessibility in FMCs and FCs, as well as BNC2 regulation of FC marker *FNDC1* through intronic and intergenic enhancers (**Fig. 7f,g**). *In silico* perturbation of BNC2 predicted a shift from a FC state toward a FMC state together with reduced expression of *IBSP*, *RUNX2*, *COL12A1*, and *FNDC1* (**Fig. 7h**). This was supported by increased *BNC2* expression in intraplaque regions of advanced coronary lesions (**Fig. 7i**). These findings suggest that *BNC2* may mediate CAD risk through osteochondrogenic SMC programs. In ECs, *CRTAC1* was predicted to be regulated by FOXC1 through intronic and intergenic regions, consistent with its motif accessibility and expression profile (**Supplementary Fig. 14**).

### Refinement of cell state-specific regulatory mechanisms at CAD GWAS loci

To prioritize candidate causal variants and target genes at CAD GWAS loci, we analyzed the publicly available MVP CAD credible set (9,437 variants)^7^ **(Methods and Supplementary Table 14)**. Most credible variants mapped to intronic, promoter and intergenic regions within L1 DARs, predicted enhancer-gene links and H3K27ac-marked active enhancers **(Fig. 8a and Supplementary Fig. 15a)**. We then partitioned these variants across L1 cell type-specific enhancers, which revealed 135 DARs in SMCs and 45 in ECs overlapping credible variants and predicted E2G enhancers **(Fig. 8b and Supplementary Fig. 15b)**. Intersecting these loci with our nominated CAD effector genes **(Fig. 7a)** highlighted known SMC CAD loci such as *PRDM16*^8^ in addition to a less defined locus such as *MAP3K7CL*. In ECs, this analysis prioritized canonical GWAS loci such as *NOS3* and *CLDN5* and less characterized loci such as *MCF2L* and *FGD5*.

**Figure 8.**
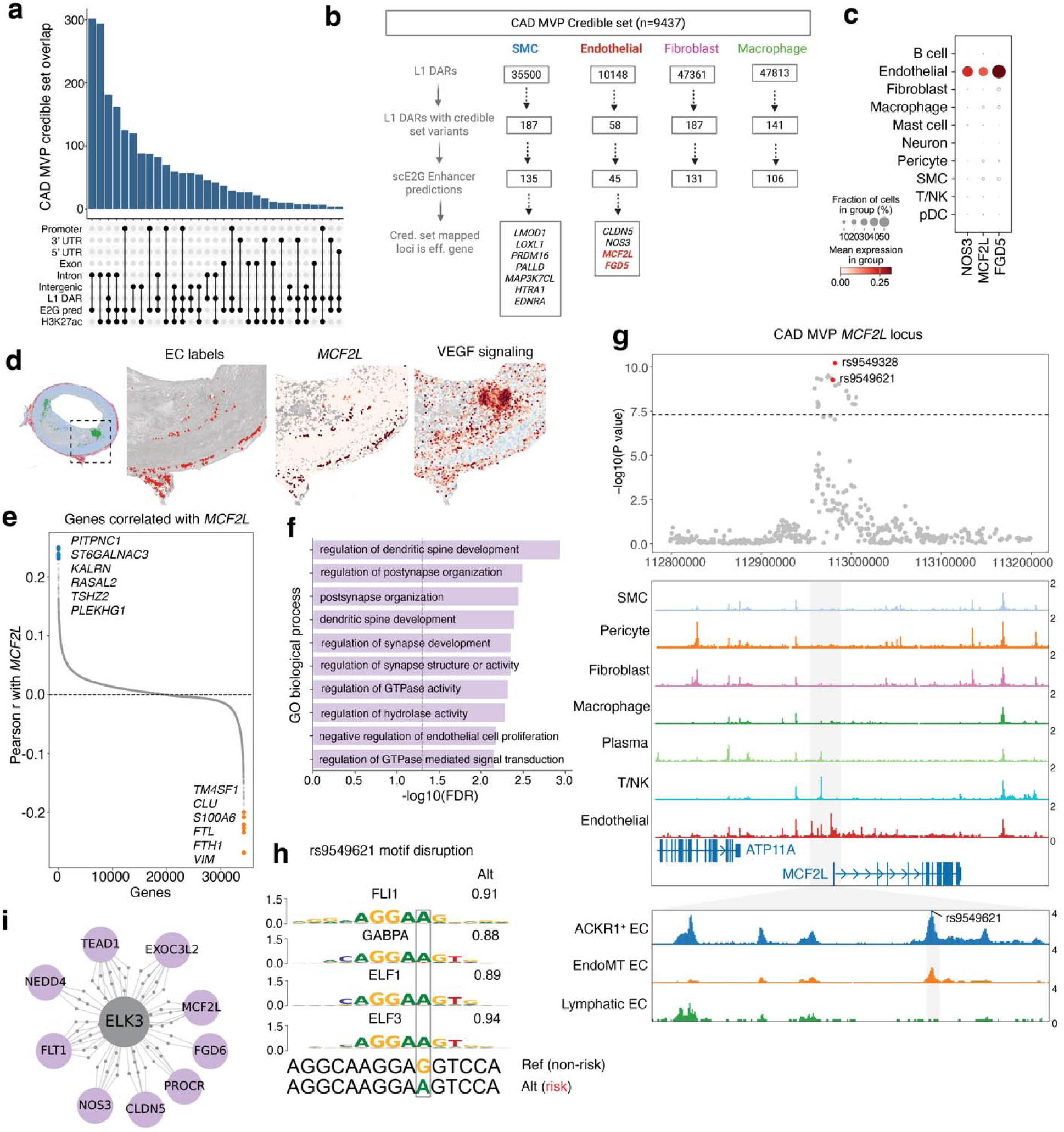
Refinement of cell state-specific regulatory mechanisms at CAD GWAS loci. **a,** UpSet plot showing categorization of fine-mapped GWAS variants located in various genomic regions. Categories include genomic annotations (e.g., promoter, exons, introns, intergenic, etc) in addition to differentially accessible regions (DARs), scE2G predicted enhancers and H3K27ac peaks. Overlaps were computed using a 100bp window for SNPs in the credible set (+/- 50 bp from the SNP coordinate). **b,** Workflow for prioritizing CAD GWAS credible set variants using cell-specific open chromatin regions. Variants (100bp windows) were first overlapped with level 1 differentially accessible regions (L1 DARs), and subsequently with scE2G predicted enhancers. Loci mapped to prioritized credible set variants were intersected with SMC and EC-specific GWAS effector genes for further downstream analyses. **c,** Dotplot showing normalized expression of prioritized Endothelial loci in **b** (*NOS3*, *MCF2L*, *FGD5*) across L1 annotations from the scRNA-seq data. **d,** Spatial plot of an advanced coronary lesion colored by L1 annotations. Insets show localization of Endothelial cells within a region of interest (ROI; left), normalized expression of *MCF2L (*middle) and inferred VEGF signaling activity (right) from Progeny. **e,** Scatter plot showing transcriptome-wide Pearson correlations of *MCF2L* in L1 Endothelial cells (ECs) from the scRNA-seq reference. The top six genes with the highest and lowest correlations, colored in blue and orange, respectively. **f,** Bar plot showing gene ontology biological processes (GO BP) enriched among the top 50 genes positively correlated with *MCF2L* as shown in **e**. **g,** LocusZoom plot showing the CAD lead SNP and prioritized credible set variant (rs9549621) from **b** at the *MCF2L* locus, both colored in red (top). Genome browser tracks showing the two variants overlap a region specifically accessible in the *MCF2L* promoter of ECs (middle). Inset shows genome tracks showing accessibility across EC L2 annotations at the *MCF2L* promoter harboring the rs9549621 SNP. **h,** Credible set variant rs9549621 was predicted to influence binding of multiple ETS family factors (FLI1, ELF3). The alternate allele A creates a binding site for ETS factors with allele diff scores > 1. The alleleDiff scores were calculated by motifbreakR with the method = “ic” and threshold=1e-4 using motif databases described in **Methods**. **i,** SCENIC+ identified network for ELK3 activating regulons in EC L1 annotations. Transcription factors shown as the central grey node, target regions represented as small grey dots and target genes shown as labeled, colored dots. EC L1 effector genes shown in purple.

*MCF2L* and *FGD5* encode Rho guanine nucleotide exchange factors (RhoGEFs) that activate Rho GTPases essential for controlling cytoskeletal dynamics during proliferative processes such as angiogenesis^57^. Consistent with our ESμ results, both genes were expressed almost exclusively in ECs **(Fig. 8c)** and localized to the adventitial layer of subclinical lesions and the adventitia and plaque of advanced HCA lesions **(Fig. 8d and Supplementary Fig. 15c)**.

Intriguingly, the *MCF2L+* region within the plaque also showed strong vascular endothelial growth factor (VEGF) signaling activity **(Fig. 8d, Methods)**. Altogether, this suggests a potential link between *MCF2L* activity in the vasa vasorum and intraplaque neovessel formation.

Rare *MCF2L* variants have been previously linked to premature atherosclerosis through impaired Rac1 and RhoA activation ^58^. To define pathways associated with *MCF2L*, we conducted a correlation analysis of this gene’s expression along the entire EC transcriptome and identified moderate correlations with CAD-relevant GEFs like *KALRN*^59^ **(Fig. 8e)**, and enrichment for regulation of GTPase activity and vascular endothelial cell function **(Fig. 8f and Supplementary Fig. 15d)**, supporting a role in angiogenic programs in human atherosclerosis. To refine candidate causal mechanisms at this locus, we mapped the sentinel SNP rs9549328 and a high-LD prioritized variant, rs9549621, to the promoter of the predominant arterial *MCF2L* transcript in GTEx **(Supplementary Fig. 15e),** within a region selectively accessible in ECs **(Fig. 8g).** L2 profiles showed that this signal was driven primarily by the *ACKR1*+ EC state **(Fig. 8h)**. Importantly, rs9549621 was replicated among multiple CAD credible sets^6,7^ and was predicted to alter ETS-family factors binding, including FLI1 and ELF1/3 **(Fig. 8h and Supplementary Fig 15f),** with the alternate allele creating a putative ETS-binding site. This regulatory link was confirmed through our eGRNs, which showed *MCF2L* as a downstream target of the ETS factor ELK3 **(Fig. 8i)**.

## Discussion

We present MetaPlaq, a multimodal single-cell atlas of human atherosclerosis, comprising more than ∼1 million cells across transcriptomics, epigenomics and high-resolution spatial modalities. Building on our previous scRNA-seq atlas^9^, our hierarchical framework harmonizes 14 datasets spanning 216 samples across different scRNA technologies and arterial beds. MetaPlaq identifies major cell lineages and more than 50 subtypes, expanding fibroblasts, pericytes and immune cells in comparison to previous atlases^9,11,12^. These data represent the most comprehensive survey of vascular and immune heterogeneity in human atherosclerosis to date, and the largest spatially resolved atlas of coronary artery transcriptomes. Spatial mapping assigned broad and granular cell states within their native tissue environment, resolving processes such as the progressive loss of SMC differentiated state upon migration from the medial layer. Integration with GWAS summary data further supports previous work showing that SMCs and ECs capture a substantial component of CAD heritability.

Using a robust expression specificity framework, MetaPlaq revealed dynamic SMC and EC gene programs during disease progression. In SMCs, osteogenic signatures were increased in advanced lesions, and most upregulated DEGs were shared across ECM-associated phenotypes rather than restricted to a single state. These findings emphasize the need for more temporal datasets to clearly distinguish programs shared along the Contractile-FMC-FC axis from those that confer more specialized functions^10^. In ECs, ECM-remodeling and angiogenic responses were upregulated, whereas downregulated genes included differentiated and immune markers, potentially reflecting early endothelial dysfunction and activation. This is consistent with increased EndoMT EC responses in coronary lesions with intraplaque hemorrhage^60^, and with assignment of upregulated genes to EC L2 states.

We also provide a comprehensive cis-regulatory landscape at single-cell resolution. More than 500,000 open chromatin regions (OCR) defined cell-specific CRE landscapes across disease-relevant cell states, approximately doubling the number of OCRs previously detected^8,17^.

Integration of advanced coronary lesion snATAC-seq data with public datasets delineated key regulators along the SMC-FMC-FC axis and three major populations of ECs, including lymphatic, *ACKR1^+^*, and EndoMT states. Guided by our expression specificity analyses, these enhancer landscapes nominated markers such as *FNDC1* and *ANPEP* for FCs and *OGN* for EndoMT, and enabled deconvolution of CAD heritability across granular states. FMC-specific CREs contributed the largest to CAD heritability among SMCs, whereas EndoMT and *ACKR1+* ECs accounted for most of the EC signal, potentially reflecting acquisition of mesenchymal programs shared with contractile SMCs and FMCs.

MetaPlaq further prioritized gene regulatory networks (GRNs) associated with CAD. Using SCENIC+^54^, we inferred enhancer-driven GRNs by harmonizing transcriptomic and epigenomic profiles. Integration with DEGs and GWAS effector genes prioritized regulons driven by TFs such as BNC2, recently associated with CAD through a multi-trait GWAS^51^. BNC2 showed increased motif accessibility and enhancer burden in FCs, with predicted downstream targeting of osteogenic markers including *IBSP* and *FNDC1.* These results suggest that BNC2-associated genetic risk may be mediated in part through osteogenic SMC programs, although its expression in multiple cell types suggests additional influences on CAD risk.

To resolve cell-specific regulatory mechanisms at CAD risk loci, we fine-mapped CAD associations using a multimodal approach integrating cell-specific effector genes with DAR-based credible set variant overlaps. This approach highlighted *MCF2L* and *FGD5* as loci that may mediate CAD risk primarily through ECs. *MCF2L* was highly EC specific and enriched in pro-angiogenic ECs localized to adventitial and intraplaque regions. *MCF2L* candidate causal variants mapped to a promoter preferentially accessible in *ACKR1+* ECs where they are predicted to disrupt ETS factor binding including FLI1, a regulator of angiogenesis^61^. These findings link *MCF2L* to neovascularization and nominate a putative regulatory mechanism at this locus. Prior studies have implicated RhoGEFs in arteries and VSMCs in hypertension^62^, further supporting *MCF2L* as a compelling candidate for functional follow-up.

A few limitations in our study should be considered. We acknowledge that the disease-stage definitions were not uniformly supported by available histology data across public datasets, limiting resolution of markers of atherosclerosis progression in differential expression analyses. Also, Visium HD is known to be sensitive to tissue quality; necrotic and fibrotic regions limit transcript detection, while bin-based capture requires post-processing for single-cell assignment, potentially introducing segmentation-based contamination. For the epigenomics data, incomplete matching of L2 SMC and EC annotations likely reflects differences in cell-type proportions across modalities. For example, the *ACKR1*+ cluster may include A*CKR1+*, pro-angiogenic, and intimal EC states identified by scRNA-seq. Expanding the snATAC reference should improve the balance and detection of underrepresented states. *In-silico* perturbations were limited to SMCs and future studies should extend L2 regulon perturbations to additional disease-relevant populations.

In summary, MetaPlaq provides a comprehensive multimodal reference for human atherosclerosis and a framework for annotating disease-associated variants and regulatory mechanisms underlying candidate causal loci. Integration of transcriptomic and epigenomic data prioritized target genes, affected cell types and regulatory programs at CAD GWAS loci with resolution of granular cell states. Future annotations from the community will be incorporated into updated atlas releases. We also provide utilities for raw data curation, and modular benchmarking workflows extending frameworks such as scIB, and our scArches compatible integration strategy^18^ is generalizable to other tissue contexts. Overall, MetaPlaq establishes a high-resolution and scalable reference for interrogating vascular and immune cell diversity, regulatory programs and disease states in human atherosclerosis.

## Acknowledgements

C.L.M. acknowledges grant support from the National Institutes of Health (NIH) (R01 HL148239, R01 HL164577, R01 HL172888, R01 HL162259, R01 HL170024, R01 HL175148, R01 AI177341, and U01 DK142283), Leducq Foundation ‘PlaqOmics’ (18CVD02) and ‘COMET’ (24CVD02) networks, Chan Zuckerberg Initiative Data Insights grant ‘MetaPlaq”, American Heart Association (AHA) Transformational Project Award (24TPA1300556) and EU Horizon Europe ‘NextGen’ grant (101136962). S.W.vdL. acknowledges grant support from EU H2020 TO_AITION (grant number: 848146), EU HORIZON NextGen (grant number: 101136962), EU HORIZON MIRACLE (grant number: 101115381), Dutch Heart Foundation ‘AtheroNETH’, NIH-NIDDK ‘Genetics of Atherosclerosis’ (grant number: 1U01DK142283-01), Health∼Holland PPP Allowance ‘Getting the Perfect Image’, and ‘SugarPlaque’ supported by EFSD and Lilly European Diabetes Research Programme. The AtheroNETH consortium is funded by the Dutch Heart Foundation (01-001-2024-0601); this collaboration is supported by the Dutch CardioVascular Alliance (DCVA). We are thankful for the support of the Leducq Fondation ‘PlaqOmics’ (18CDV-02) and ‘AtheroGen’, and the Chan Zuckerberg Initiative ‘MetaPlaq’. The research for this contribution was made possible by the AI for Health working group of the EWUU alliance (https://aiforhealth.ewuu.nl/). The collaborative project ‘Getting the Perfect Image’ was co-financed through use of PPP Allowance awarded by Health∼Holland, Top Sector Life Sciences & Health, to stimulate public-private partnerships. A.Y.Jr. acknowledges grant support from NIH: R01 HL182859, R01 HL167758, R01 HL182756, R01 HL180481, and the U.S. National Science Foundation (2537597).

We acknowledge support from the University of Virginia School of Medicine Spatial Biology Core Facility, RRID: SCR_023281. We also thank Dr. Pat Pramoonjago and Angela Miller in the UVA Biorepository and Tissue Research Facility (BTRF) as well as Cristie Liu for histological processing and staining. Biorender was used to create the graphical abstract in Fig. 1 and the schematics in Fig. 5a, Fig. 5j, and Fig. 8b.

## Author Contributions

C.L.M. supervised research primarily related to the study. N.J.L., A.V.F., J.C.K., J.L.M.B., and S.W.vdL. jointly supervised research secondarily related to the study. J.V.M., M.M., A.Y.Jr., S.W.vdL, and C.L.M. conceived and designed the experiments. J.V.M., G.A., M.L., A.K.dO., K.S-C., and A.W.T. performed the experiments. J.V.M., I.T., and M.M. performed the statistical analyses. J.V.M., I.T., M.M., A.K., P.H., D.M.S., A.W.T., C.J.H., K.T., T.J., and K.I.M.vdS. analyzed the data. S.S.A., R.B.C., F.V., J.C.W., P.C., C.G., N.J.L., A.V.F., J.C.K., J.L.M.B., A.Y.Jr., and S.W.vdL. contributed data/resources/analysis tools. J.V.M., I.T., M.M., G.A., S.W.vdL. and C.L.M. wrote the paper.

## Disclosures

C.L.M. has received grant support from AstraZeneca and Johnson & Johnson for unrelated work and serves on the advisory board for Vascentis. S.W.vdL. has received Roche funding for unrelated work and is a co-founder of AtheroSCREEN. Roche nor AtheroSCREEN have had no part in this study, neither in the conception, design and execution of this study, nor in the preparation and contents of this manuscript. All other authors have declared no competing interests.

## Methods

### Ethics statement

Collection of human coronary artery tissue samples for bulk RNA-seq, snATAC-seq, Visium HD, as well as histology described in this manuscript complies with ethical guidelines for human subjects research under approved Institutional Review Board (IRB) protocols at Stanford University (no. 4237) and the University of Virginia (no. 210164 and no. 302407), for the procurement and use of human tissues and information. All studies were conducted with the Declaration of Helsinki, written informed consent was obtained from all participants, and unless otherwise noted, local medical ethics or review boards approved those studies. Additional details about age, sex, and ancestry for each individual have been previously described in Hodonsky et al^63^. Ethical statements for all other participating studies are included in Supplementary Tables with links to the original manuscripts and available datasets.

#### Coronary artery tissue procurement in COSMOS

Freshly explanted hearts from orthotopic heart transplantation recipients were procured at Stanford University and the University of Virginia under approved IRB protocols and with written, informed consent. Briefly, hearts were arrested in cardioplegic solution and rapidly transported from the operating room to the adjacent laboratory on ice. The proximal 5–6 cm of three major coronary vessels (left anterior descending, left circumflex and right coronary artery) were dissected from the epicardium on ice, trimmed of surrounding adipose and rinsed in cold PBS and rapidly snap-frozen in liquid nitrogen. Coronary artery samples were also obtained at Stanford University (from Donor Network West and California Transplant Donor Network) from nondiseased donor hearts rejected by surgeons for heart transplantation and procured for research studies. All hearts were procured after written informed consent from legal next-of-kin or authorized parties for the donors. Reasons for rejected hearts include size incompatibility, comorbidities or risks for cardiotoxicity. Donor hearts were arrested in cardioplegic solution and transported on ice following the same protocol as explanted hearts. Explanted hearts were generally classified as ischemic or nonischemic cardiomyopathy and previous ischemic events and evidence of atherosclerosis was obtained through retrospective review of electronic health records at Stanford Hospital and Clinics. The disease status of coronary segments from donor and explanted hearts was also evaluated by gross inspection at the time of harvest (for presence of lesions), as well as histological analysis of adjacent frozen tissues embedded in OCT blocks. Frozen tissues were transferred to the University of Virginia through a material transfer agreement and IRB-approved protocols. All samples were stored at −80L°C until the day of processing. Tissues were de-identified and clinical and histopathology information was used to classify ischemic, non-ischemic hearts and lesion- and non-lesion-containing arteries. All normal arteries originated from hearts with left ventricular ejection fraction greater than 50%.

#### Carotid artery sample processing and library preparation

The detailed procedures describing the processing of plaques, isolation of RNA, and CEL-Seq2 experiment from the AtheroExpress biobank study (https://atheroexpress.nl/) have been described before^64^. In short, plaques were processed within 10 minutes of surgical removal. A 5 mm culprit segment was preserved in paraffin for histology; the remainder was minced and enzymatically digested in RPMI 1640 containing Collagenase IV (2.5 mg/mL), DNAse I (0.25 mg/mL), Human Albumin Fraction V (2.5 mg/mL), and Flavopiridol (1 mM) at 37°C for 30 minutes. The resulting suspension was filtered through a 70 µm strainer. Viable cells were isolated by FACS (Beckman Coulter MoFlo Astrios EQ) using Calcein AM and Hoechst staining; remaining unstained cells were cryopreserved. scRNA-seq was performed using the SORT-seq protocol with CEL-Seq2 library preparation. Single viable cells were sorted into 384-well plates pre-loaded with lysis buffer containing CEL-Seq2 primers, spike-ins, and dNTPs, and immediately frozen at −80°C. After cDNA synthesis and *in vitro* transcription, libraries were sequenced paired end (2×75 bp) on an Illumina NextSeq 500.

### MetaPlaq atlas construction

#### scRNA-seq raw sequencing alignment

FASTQ files from 10X Genomics datasets were aligned using Cell Ranger (v.8.0.0) using the 10X hg38 genome reference (v2024) downloaded at the following link **wget** "https://cf.10xgenomics.com/supp/cell-exp/refdata-gex-GRCh38-2024-A.tar.gz".

Smart-seq2 datasets from Mocci et al^65^ were processed with a custom pipeline wrapping STARsolo^66^ (v.2.7.9) with the following parameters –soloType=SmartSeq –soloUMIdedup Exact –soloCellFilter None –outSAMattributes RG –outFilterScoreMinOverLread 0.3 – outFilterMatchNminOverLread 0.3 –soloFeatures Gene GeneFull. Raw paired-end reads from CEL-Seq2 libraries (NextSeq 500, 2×75 bp) were processed using the celseq2 pipeline (https://github.com/yanailab/celseq2) within a dedicated mamba environment on a SLURM-managed HPC. For each sample, lane-split FASTQ files (L001–L004) were concatenated per read using cat to produce merged R1 and R2 files compatible with the celseq2 experiment table format. Reads were then processed using celseq2-slim with a project-specific configuration file, performing demultiplexing of the 384 cell barcodes per plate, UMI extraction, and alignment to the GRCh38 reference (10x Genomics refdata-gex-GRCh38-2024-A) via the bundled aligner.

Resulting SAM files were converted to BAM, sorted with samtools, and merged per sample using custom Python wrappers. Per-sample expression count tables, annotation files, and run reports were aggregated into a unified output structure for downstream quality control and integration. This pipeline was applied to 39 samples, comprising 64 carotid plaque libraries from the Athero-Express Biobank Study, generating the count matrices used as input to the QC and integration workflow described below.

#### Quality control for individual scRNA libraries

Feature count matrices generated by the above alignment pipelines were processed using a QC pipeline implemented in our scRNAutils R package (https://github.com/MillerLab-CPHG/scRNAutils/tree/dev). Within this pipeline, we first identify doublets for each library by performing 3 iterations of artificial doublet generation and detection with scDblFinder^67^ (v.1.16.0). Consensus doublets across all iterations were removed from downstream analyses. Ambient RNA contamination is a key issue as it can lead to cross-contamination of gene expression and confound clustering and marker identification. To correct for ambient RNA, we ran DecontX^68^ within the celda R package (v.1.18.2) in doublets-filtered raw counts matrices using default parameters. Decontaminated raw count matrices output by DecontX were then added into each Seurat object^19^.

Given the high variability in terms of quality and sequencing depth across the included datasets, we opted for further filtering using an adaptive thresholding approach implemented in our pipeline. First, we build a full rank matrix of cells by QC covariates (% Mitochondria, %Ribosomal reads, N Reads and N Genes). We then compute an outlyingness score using the robustbase R package (v0.99.7) with the **adjoutlyingness** function. Each cell receives an outlyingness score and then we remove outliers based on the Median Absolute Deviation (MAD) using the **isOutlier** function implemented within the scater R package^69^ (v1.30.1). We chose a rather stringent MAD threshold since higher MADs led to an increased proportion of low-quality cells. Because this feature is quite sparse, we also remove cells with > 1% reads mapped to hemoglobin genes as these are likely contaminating erythrocytes.

Each counts matrix was then normalized using the **SCTransform** function within the glmGamPoi R package^70^ (v.1.14.3) with *vst.flavor = v2*, which corrects for technical confounders in measured gene expression such as per-cell sequencing depth. For each library, technical factors such as cell cycle and mitochondrial read percentage were regressed out during SCTransform normalization. After normalization, we curated a set of 3000 highly variable genes (HVGs), removing MALAT1 and mitochondrial/ribosomal genes to prevent artifacts and housekeeping genes from biasing dimensionality reduction. We then carried out dimensionality reduction using principal component analysis (PCA) using the **RunPCA** function from the Seurat package^19^ (v.5.0.1) and selected the number of PCs explaining 90% of variance in the data using a custom function. PCA embeddings were then used for creating a shared nearest neighbors graph with the **FindNeighbors** function from Seurat. To find the proper clustering configuration for each library, we created an Euclidean cell low-dimensional space using the **dist** function from the stats R package (v.4.3.1) and clustered the data using the **findClusters** function across a range of resolutions (0.3-1). We then used the **silhouette** function from the cluster R package (v.2.1.8.2) to compute silhouette scores for each clustering resolution.

#### Integration benchmarking

Raw pseudocounts (post-ambient RNA correction) were concatenated using the anndata package (v.0.10.9) with setting the parameter *how=”outer”* according to scverse guidelines. Upon concatenating the data, counts were normalized using the scanpy^71^(v1.9.6) package preprocessing module with the functions **scanpy.pp.normalize_total** and **scanpy.pp.log1p**. Highly variable genes were detected using the **scanpy.pp.high_variable_genes** function (*flavor=”seurat_v3”*, *n_top_genes=3000*, *batch_key=”sample”*). Dimensionality reduction with PCA was performed using the **scanpy.tl.pca** function from the tools module and a k-Nearest-Neighbors (kNN) graph built using **scanpy.pp.neighbors** with the number of neighbors set to the default.

To find the optimal approach for data harmonization, we evaluated linear embedding and deep-learning-based approaches using an in-house framework (MetaPlaq-benchmarker). In addition to leveraging GPU-accelerated metrics from the scIB package^72^ (v.0.5.1), such as Local Inverse Simpson’s Index (LISI) scores, we provide custom implementations of metrics such as batch entropy and kNN overlap (kNN purity). We aimed to implement a modular approach by creating functions that calculate metrics using a single embedding as input. We then implement wrappers that can take a dictionary of embeddings for comparing multiple batch correction approaches at once. Our Python package implements the following metrics across two criteria:

Conservation of Biological signal:

- Cell local inverse Simpson Index (cLISI).
- K-nearest-neighbors overlap (kNN purity).

Batch mixing:

- Average silhouette width (ASW).
- Integration local inverse Simpson Index (iLISI).
- Batch entropy.

We used the above metrics to evaluate the following integration algorithms:

#### Seurat-based integration

We performed batch correction with the following Seurat-supported^73^ algorithms: Canonical Correlation Analysis (CCA)(v.5.1.0), reciprocal PCA (rPCA)(v.5.1.0), and Harmony (v.1.2.4). The following workflow was applied to run each method: a list of processed Seurat objects was created, then merged, and the 3000 most highly variable genes was extracted using **SelectIntegrationFeatures**. We then ran PCA using highly variable genes with the function **RunPCA** (*npcs=10*). We then integrated datasets using the **IntegrateLayers** function and set the appropriate corresponding class for the *method* parameter:

- CCA → *method*=CCAIntegration
- rPCA → *method*=RPCAIntegration
- Harmony → *method*=HarmonyIntegration

Other parameters for **IntegrateLayers** included *normalization.method=”SCT”* and *k.param=20* using the computed PCs in the prior step. The batch-corrected embeddings were supplied to compute a shared-nearest-neighbors (SNN) graph using 10 PCs with the function **FindNeighbors**. Louvain clustering was then performed with the function **FindClusters** (*resolution=2*), and UMAP embeddings were computed with the **RunUMAP** function.

### STACAS

The STACAS package^22^ (v.2.2.2) implements a batch correction using mutual nearest neighbors to identify biologically equivalent cells (“anchors”) across dataset pairs. Building on Seurat integration, STACAS uses reciprocal principal component analysis (rPCA), projecting each dataset into the principal component (PC) space of the other in the pair to identify anchors and weight their biological relevance based on rPCA distance for batch correction. To use this workflow, we first created a list of processed Seurat objects. Batch correction was performed with the function **Run.STACAS (***min.sample.size=0, dims=10, k.weight=45*). This function also calculates nearest neighbors after integration. Finally, UMAP embeddings were computed using the Seurat **RunUMAP** function.

### Harmonypy

The Harmony algorithm^20^ corrects batch effects in integrated scRNA-seq datasets by iteratively adjusting low-dimensional cell embeddings to maximize mixing across user-supplied batch labels. Processed Seurat objects were converted to anndata using sceasy (v.0.0.7). Anndata objects were merged using the ***concat*** function from the anndata Python package. The merged anndata object was normalized using the **scanpy.pp.normalize** and **scanpy.pp.log1p** functions. PCA embeddings were obtained with the function **scanpy..tl.pca function** after computing 3000 HVGs with the **scanpy.pp.highly_variable_genes** function. Batch effects were corrected using the harmonypy package^20^ (v.0.0.10) with the function **harmonypy.run_harmony** and added back into the merged anndata object. Harmony embeddings were then used for computing an n-nearest neighbors graph with the function **scanpy.pp.neighbors** and subsequently used for Leiden clustering with the function **scanpy.tl.leiden** setting *n_iterations=2*.

### Scanorama

Scanorama^21^ leverages a mutual nearest neighbors (MNN) approach to correct for batch effects by identifying shared cell populations in a low-dimensional embedding space. It then computes translation vectors between matched cell populations to align datasets to preserve dataset-specific biological variation. To use this algorithm, we first created a list of processed anndatas and converted each counts matrix to a .csr object using the **csr_matrix** from the scipy Python package. To integrate datasets, we then used the **correct_scanpy** function from the scanorama package^21^ (v.1.7.4) setting *return_dimred=True*. We then created a concatenated object containing batch-corrected embeddings and corrected count matrices using the anndata **concat** function setting *join=”outer”*. After obtaining a single batch corrected object, we computed a nearest-neighhbors graph and performed Leiden clustering as done with the harmonypy approach.

### Single-cell Variational Inference (scVI)

The single-cell Variational Inference approach (scVI)^26^ package leverages a variational autoencoder (VAE) framework to create a low-dimensional embedding of single-cell gene expression data. By conditioning the decoder on batch labels, scVI generates an embedding that minimizes technical variation due to batch effects while preserving biologically meaningful transcriptional differences. To use this framework, 3000 HVGs were computed as detailed above setting *flavor=”seurat_v3”*. Setup before model instance creation and training was done using the **SCVI.setup_anndata** function from the scvitools package^26^ (v.1.1.5) setting *layer=”counts”* and *batch_key=”sample”*. We tested different training epochs and selected 100 based on observed value updates of the loss function. We then created a scVI model instance using the **model.SCVI** class (*n_hidden=128, n_latent=10, n_layers=2, gene_likelihood=”zinb”*) and trained for the above number of epochs. The scVI latent space was then extracted with **model.get_latent_representation** and used for nearest neighbors computation and Leiden clustering with the functions **scanpy.pp.neighbors** and **scanpy.tl.leiden** before obtaining UMAP embeddings with **scanpy.tl.umap.**

### Single cell architectural surgery (scArches)

The architectural surgery (scArches)^18^ workflow is a transfer learning approach that takes an existing reference model (e.g., variational autoencoder; VAE) and adapts it to enable query-to-reference mapping. After training an existing autoencoder model on a reference dataset, architectural surgery is the process of transferring these trained weights with only minor weight adaptation (fine-tuning) and adding a condition node to map new studies into the reference. To employ this approach, we performed data normalization as detailed above and selected 3000 HVGs setting *flavor=”seurat_v3”*, and *n_top_genes=3000*. We then used raw counts in our reference to create a scVI model (*encode_covariates=True, dropout_rate=0.2 and n_layers=2*), which was then trained setting *max_epochs=100*. This scVI baseline model was used to train a reference with a semi-supervised scANVI^74^ model with the following parameters (*max_epochs=20, n_samples_per_label=100*) and level 1 (L1) labels from our previous atlas^9^. We prepared our query data for weight fine-tuning of the reference using the **scvi.model.SCANVI.prepare_query_anndata** and **scvi.model.SCANVI.load_query_data** functions. Given the complex batch effects observed in the data, the query model was trained by iterating across multiple epochs (100, 150 and 200). The final parameters used for training the query model were as follows (*max_epochs=200, plan_kwargs={“weight_decay”:0.0}, check_val_every_n_epoch=10*). The scArches latent space was used for computing UMAP embeddings with scanpy. L1 cell annotation labels were then transferred to the new reference using the query model **predict** method setting *soft=True*.

#### Post-integration Quality Control and Level 1 annotations

After QC and harmonization, we used the scArches^18^ latent space for computing a kNN graph with the function **scanpy.pp.neighbors** setting n_neighbors=15. This graph was used as input for cell clustering with the Leiden algorithm, iterating across the range of resolutions (1.5-1.8) based on the size of the newly-created reference. A resolution of 1.5 was chosen as it distinguished major vascular and immune clusters without overclustering the data. We then performed a first pass of manual annotations based on canonical markers. Then, we refined labels by cross-referencing our annotations against predicted labels. We flagged clusters that showed a mixture of predicted labels, were dispersed across multiple lineages and/or contained abnormally low prediction confidence scores as these likely depicted remaining doublets. Upon removing low-quality clusters, we did a second pass of annotations to establish the final L1 labels.

#### Level 2 annotations

To add level 2 (L2) annotations, each L1 compartment was subset. After subsetting, we performed a second round of HVGs (n=3000) detection to find meaningful genes for a given L1 compartment. We used L1-specific HVGs to train a scVI model for each L1 cell type separately using the following parameters (n_hidden=128, n_layers=2, latent_distribution=”normal”, dispersion=”gene”, n_latent=30 and gene_likelihood=”nb”). We then used the embeddings generated by scVI to perform a new round of Leiden clustering across a given range of resolutions. We selected the resolution that better delineated the expression of known markers in order to add level 2 (L2) annotations. The scVI latent space was processed as described above to obtain UMAP embeddings for each L2 annotation. For each compartment, we performed Leiden clustering across several ranges of resolutions determined based on the size of the L1 compartment. Finally, we chose a resolution that delineated the expression of markers described in our previous work^9^ and avoided data overclustering. The final set of L2 annotations was added based on our previous set of markers in addition to a literature search. Cells from a lineage other than the target L1 compartment were labeled as “stripped cells” and removed from further downstream analyses.

#### Enrichment of SMC murine gene signatures and TF regulon targets

Given the intrinsic phenotypic plasticity of SMCs, we validated our manual annotations using gene set enrichment with murine lineage-traced SMC gene sets. Marker tables for each annotated SMC state were extracted from *Li et al 2026*^10^. Markers for each SMC phenotype were ranked by Log2FC value and then converted to human nomenclature using a custom script that leverages the biomaRt database (v.2.58.0). We refined feature sets by keeping only genes with a one-to-one ortholog relationship for downstream enrichment. Curated gene sets were used as input for single–cell resolution gene set enrichment within the SMC compartment using the decoupler^28^ (v.2.1.1) **dc.mt.ulm** function with the parameter *tmin=3*.

For enrichment of TF regulon targets with single cell resolution, we first loaded the CollecTRI^75^ collection of human regulons using the **collectri** function from decoupler^28^. Regulons were then scored across L1 and L2 datasets using the **dc.mt.ulm** function as described above. Marker TFs were identified using the **rankby_group** function from decoupler with the parameters *reference=”rest”* and *method=”t-test_overdistim_var”*.

#### Collection and pre-processing of GWAS summary statistics

Curating these GWAS summary statistics at scale exposed a range of practical obstacles. Not all datasets are publicly available; some require author contact, and others only report lead variants. Through private communications with corresponding authors or through public download we collected GWAS summary statistics for: CAD from Million Veterans Program (MVP)^7^ (N sample size = 773,268); Myocardial infarction^45^ (N sample size = 471,717); Coronary Artery Calcification (CAC) multi-ancestry GWAS meta-analysis^38^(N sample size = 26,909), carotid intima-media thickness (cIIMT) and carotid plaque presence (PP)^76^ (N sample size = 48,434; N significant loci = 5); any stroke^77^ (N sample size = 1,308,460); any ischemic stroke^77^ (1,296,908); pulse pressure, diastolic blood pressure and systolic blood pressure from the Million Veterans Program^44^ (N sample size = 318,890); Alzheimer disease^78^ (N sample size = 455,258) type 2 diabetes (UK Biobank)^79^ (N sample size = 455,607); LDL and HDL cholesterol (N sample size = 1,320,016); body mass index (UK Biobank) (N sample size = 457,824)^79^.

Details for these GWAS summary statistics datasets are available in Supplemental Table 11. Beyond access, inconsistencies in reported sample size, genome build, and missing fields — particularly standard errors and effect sizes — were common. To handle this systematically, we developed gwas2cojo (https://github.com/CirculatoryHealth/gwas2cojo), a custom Python pipeline building on gwaslab^80^, which automates genome build liftover, allele alignment against a population reference, and output to a uniform file format. The pipeline was first described in Slenders et al.^81^; the version applied here incorporates several extensions described below.

Study selection followed a straightforward hierarchy: phenotypic relevance, the largest and most recent available study, and preference for open-access or easily downloadable data. Summary statistics with incomplete critical fields which could not be computed from the available fields were excluded at this stage.

Pre-processing and primary quality control were performed using gwas2cojo’s GWASLab Processing pipeline **(gwas_process.py)**. All genomic positions were lifted over to GRCh38/hg38 where required - using hg18->hg38 or hg19->hg38 chain files as appropriate - and alles were aligned against the 1000G Genomes Project phase 3 v5 reference (hg19) studies or the 1000 Genomes 30x reference (hg38 studies). Effect allele frequencies (EAF) absent from the source files were retrieved from the matched population 1000 Genomes reference by direct position (CHR:POS) and allele lookup. rsID assignment used dbSNP build 151. Studies reporting odds ratios rather than log-scale effect sizes had betas derived automatically as β = ln(OR) prior to ingestion. When standard errors were not reported these were derived as:

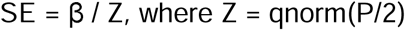

with β the reported effect size and P quantile-normalised using R’s qnorm function. Variants were retained if EAF ≥ 0.5%, imputation quality (INFO) ≥ 0.4, |BETA| ≤ 5, and SE ≤ 5.

The final set of summary statistics was organised into three thematic groups: atherosclerotic cardiovascular disease (n = 7), cardiometabolic and vascular risk factors (n = 7), and other traits (n = 1).

#### MAGMA

Similar to Slenders et al.^81^, gene-level analysis was conducted with MAGMA (v1.07), and functional annotation and prioritisation of significant loci were performed through FUMA v1.3.6 (https://fuma.ctglab.nl/). Independent loci were defined using a genome-wide significance threshold of P < 5 × 10⁻L and LD clumping at r² > 0.05 within 1,000 kb of the lead variant.

#### Single cell disease relevance scores (scDRS)

We used the single-cell disease relevance score (scDRS) framework^24^ to identify cell subtypes driving the GWAS enrichments previously shown in L1 annotations. scDRS integrates gene expression profiles from scRNA-seq with polygenic disease information from GWAS to associate individual cells to disease by assessing the excess expression of GWAS putative disease genes in a given cell relative to other genes with similar expression across all cells.

For these analyses, we selected MAGMA gene sets from CAD, MI, Plaque, CAC, Plaque, PP as representative cardiovascular traits and immune traits showing enrichment of level 1 Macrophage annotations (e.g., Alzheimer’s disease) to better guide the specificity of cardiovascular enrichments. To run scDRS, we selected the top 1,000 MAGMA features from each gene set, weighted by their *P* values as putative disease gene sets. Munging of MAGMA gene sets for scDRS was performed using a custom NextFlow pipeline using a custom script and the scDRS command line interface^24^ (cli) (v.1.0.4), specifically **scdrs munge-gs** with the parameter *–n-max=1000* to select top 1000 associated genes.

We ran the enrichments in the entire meta-analyzed reference as it is recommended to use data harboring a diverse set of cells with varying relevance to the GWAS traits of interest. To prepare the count matrix for disease score enrichments, we used the **scdrs.preprocess** function with *n_mean_bin=20* and *n_var_bin=20*. Prior to the computation of normalized disease scores, we generated 1000 control gene sets matching the mean and variance of each target disease/trait gene set. Finally, we calculated normalized disease scores using the **score_cell** function.

Normalized disease scores for each cell in the reference were then added back into the meta-analyzed anndata object and visualized with scanpy.

#### Comparison of level 2 SMC and EC transcriptomic profiles

We grouped cells in SMC and EC subsets by their corresponding L2 annotation and then computed mean normalized expression across their 3000 HVGs. Cosine similarities were computed using the function **cosine_similarity** from the scikit-learn package (v.1.7.2). To measure Euclidean distances in PCA space, we first reduced the dimensionality of the L2 subset with the PCA class from scikit-learn. PCA embeddings were then used as input for the **squareform** function from the scipy package (v.1.14.1) to compute Euclidean distances.

#### Level 1 and 2 expression specificity coefficients computation

We derived expression specificity estimates for each gene across each cell annotation using the approach proposed by Timshel et al.^27^. While previous approaches for genetic prioritization of cell types have used binary or discrete representations of cell type expression^78,82^, the CELLEX workflow uses a continuous representation of cell type expression. CELLEX combines 4 different Expression Specificity (ES) metrics into a single specificity estimate. Briefly, gene expression specificity weights (ES_W_) were calculated separately for each of the following ES metrics: Gene Enrichment Score (GES)^83^, Expression Proportion (EP)^84^, Normalized Specificity Index (NSI) and Differential Expression T-statistic (DET). ES_W_ values were then averaged into a single ES estimate (ES_μ_), assuming equal weights for each metric. An ES_μ_ value represents the score that a gene is specifically expressed in each cell type. This combined metric has been shown to be more robust than single-expression specificity measures.

As a pre-processing step before computing specificity values, we extracted raw counts from L1 and L2 matrices. We summed counts for all genes across cells in each matrix and computed distribution deciles. To reduce the burden of artifacts from lowly-expressed genes, we kept genes above the third decile. Along gene-level filtering, we created a metadata matrix containing a label for each barcode (L1 or L2 annotations). Filtered raw counts and annotation metadata was used as input to create a ESObject with the cellex package^27^ (v.1.2.2) and then we computed expression specificity (ES_μ_) values using the **eso.compute** function with default settings.

#### Differential gene expression analyses

Differential expression analyses were conducted using a pseudobulking approach to account for the sparsity of scRNA-seq data. We pseudobulked raw counts from our scRNA reference across samples and Level 1 annotations using the function **pseudobulk** from the decoupler package^28^ (v2.1.2) with the parameter *mode=”sum”*. To remove low quality or low information pseudobulked profiles, we only retained those that had > 10 cells and > 1000 total counts.

Additionally, to remove artifacts from lowly expressed genes, we performed gene-level filtering in L1 cell types separately to account for intrinsic differences in expression programs across broad cell lineages. For each L1 annotation, we only retained genes that had more than had a minimum of 50 read counts across all pseudobulked profiles for a given cell type and a minimum of 10 counts in each sample. To account for lowly-expressed artifacts and multiple testing burden, we subset the L1 anndata to each main cell type annotation. For every cell type, we only retained genes that had > 50 read counts across all pseudobulked samples and >= 10 counts per sample. We also removed genes that were not present in at least 25% of cells in each pseudobulked profile.

To identify sources of variation and potential confounders, we reduced the dimensionality of the data using the PCA implementation within scanpy^71^ after normalizing and scaling raw counts as described previously. As we found that “sequencing_platform” and “tissue_source” drove most of the variance along the first 2 PCs, these were included as covariates for the Deseq2 design (∼lesion_stage + seq_platform + tissue_source). We then fit generalized linear models (GLMs) to raw counts and ran the subsequent Wald tests using the **dds.deseq2** and **DeseqStats** function from the pyDeseq2 Python package^85^ (v.0.5.3) with the parameters *alpha=0.05*, *cooks_filter=True* and *independent_filter=True*. Differentially expressed genes were designated as those with FDR < 0.05 and log2FC > 1.5 and visualized using a custom ggplot2 (v.4.0.2) script.

#### GWAS cell-specific effector genes

After computing *Esmu* values as described above, for each L1 or L2 annotation separately, we ranked genes based on their expression specificity and kept those above the 95th percentile of specificity. We also ranked genes from MAGMA files according to their *P* values and only kept the top 1000 genes per trait, consistent with the number of genes used for scDRS analyses. Finally, we notated cell-specific GWAS effector genes by intersecting the two ranked sets.

#### Bulk RNA-seq analysis in human coronary arteries

Human coronary artery samples used for bulk RNA-seq were processed as detailed previously^63^. Briefly, total RNA was extracted from frozen coronary artery segments using the Qiagen miRNeasy Mini RNA Extraction kit (catalog #217004). Approximately 50 mg of frozen tissue was pulverized using a mortar and pestle under liquid nitrogen. Tissue powder was then further homogenized in Qiazol lysis buffer using stainless steel beads in a Bullet Blender (Next Advance) homogenizer, followed by column-based purification. RNA concentration was determined using Qubit 3.0 and RNA quality was determined using Agilent 4200 TapeStation. Samples with RNA Integrity Number (RIN) greater than 5.5 and Illumina DV_200_ values greater than 75 were included for library construction. Total RNA libraries were constructed using the Illumina TruSeq Stranded Total RNA Gold kit (catalog #20020599) and barcoded using Illumina TruSeq RNA unique dual indexes (catalog # 20022371). After re-evaluating library quality using TapeStation, individually barcoded libraries were sent to Novogene for next-generation sequencing. After passing additional QC, libraries were multiplexed and subjected to paired-end 150 bp read sequencing on an Illumina NovaSeq S4 Flowcell to a median depth of 100 million total reads (>30 G) per library.

The raw passed filter sequencing reads obtained from Novogene were demultiplexed using the bcl2fastq script. The quality of the reads was assessed using FASTQC and the adapter sequences were trimmed using trimgalore. Trimmed reads were aligned to the hg38 human reference genome using STAR^66^ (v2.7.3a) according to the GATK Best Practices for RNA-seq. To increase mapping efficiency and sensitivity, novel splice junctions discovered in a first alignment pass with high stringency, were used as annotation in a second pass to permit lower stringency alignment and therefore increase sensitivity. PCR duplicates were marked using Picard and WASP^86^(v0.3.4) was used to filter reads prone to mapping bias. Total read counts and Transcripts per million (TPM) normalization was performed with the following formula after pulling exong lengths with the GenomicFeatures R package (v.1.54.4):

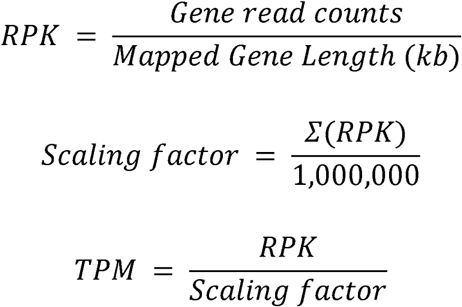

#### Coronary artery lesion stage classification

Coronary arteries were visually assessed by two independent observers and classified according to the Stary scoring system. To address limitations in statistical power due to limited sample sizes in certain assays, Stary stages were grouped into three categories: a/ early lesions, Stary stage I gathering adaptive intimal thickening or no lesions); b/ intermediate lesions, gathering Stary stages II–III defined by the presence of fatty streaks or lipid pools), and advanced lesions: Stary stages IV–VI defined by the presence of an atheroma with or without calcification/complications.

### Visium HD spatial transcriptomics analysis in coronary artery

#### Sample processing and RNA quality assessment

RNA quality of fresh-frozen coronary artery samples was evaluated using the RNA Integrity Number (RIN), which was ≥4.0. Tissue morphology was assessed with H&E and DAPI staining before Visium HD. Sections were cut to 10 μm thickness as per the Visium HD FF Tissue Preparation Handbook (CG000763, 10x Genomics), mounted on BOND 380 slides, and stored at -80°C. Slides were used within 1 week to prevent RNA degradation.

Tissue sections were stained with H&E and imaged at 20x magnification. After imaging, the coverslip was removed, and sections were destained and permeabilized with 1% SDS, then incubated for 1 hour in 70% cold methanol.

#### Visium HD library preparation

Samples were processed following the Visium CytAssist Spatial Gene Expression Reagent Kits User Guide (CG000685), with hybridization performed at 50°C for 19 hours, followed by washes and probe ligation. The transcriptomic probes were transferred onto the Visium HD slide using the CytAssist instrument for 30 minutes at 37°C to capture the probes while preserving their spatial localization. After releasing the probes from the HD slide, the captured mRNA molecules were reverse transcribed into cDNA to create libraries. These libraries were indexed with the Dual Index Kit TS Set A (PN-1000251, 10x Genomics) and purified using SPRIselect (Beckman Coulter). The sequencing depth was determined by the number of tissue spots covered. The HD slide contains a continuous array of oligonucleotides arranged into about 11 million 2 x 2 µm barcoded squares without gaps, within a 6.5 x 6.5 mm capture area, preserving the original location of mRNA in the tissue.

#### Image and read alignment

H&E-stained images were manually aligned to the Cytassist image using Loupe Browser (v.9.0.0). Reads and images were processed using Space Ranger version 4.0.1(10x Genomics)(https://www.10xgenomics.com/support/software/space-ranger/latest), reads were aligned to the Probe Set v2 (GRCh38 2020-A) and to corresponding bins, which were mapped back to the Cytassist and H&E-stained tissue images. The filtered count matrix was used for downstream processing and data analysis.

#### Quality control and segmentation

Bins with no gene expression were removed from the analysis. Samples with 4um bins were processed using Bin2Cell^25^(v.0.3.3) to perform cell grouping based on H&E image segmentation and visualization of gene expression.

#### Sample quality control

Due to a low average number of genes expressed per 4μm bin, 3 samples were excluded from a downstream analysis (samples 5, 6, 16). Genes expressed in more than 3 segmented cells were retained, and cells with minimum of 3 expressed genes were kept for downstream analysis. All the bins outside of intact coronary tissue were removed. Analysis was done using scanpy^71^(1.11.0) and anndata (v0.11.3).

#### Manual annotation of anatomical regions

If more than 1 section was present per Visium HD slide, sections were annotated based on their patient ID using our custom annotation tool that will be available upon publication.

### scVI and scANVI annotations

#### Level 1 annotations

To annotate samples, MetaPlaq RNA described in this publication was used as a reference, subsetted to the genes present in Visium HD data. 2000 HVGs were selected using scanpy (v1.12) highly_variable_genes function with flavor = “seurat” and the scVI model was trained with 2 layers and 30 latent dimensions. Then, scANVI model was created from the scVI^26^ (v1.4.2) model and trained using 20 epochs. Subsequently, the labels were transferred to the Visium HD data using the same model (*max_epochs = 100*).

### Hierarchical Level 2 annotations

MetaPlaq RNA data were subset by major cell type (SMC, Fibroblast, Endothelial, and Macrophage). For each subset, independent scVI (models were trained using Level 2 annotations. The resulting models were then used to transfer labels to the corresponding cell type–restricted Visium HD data, following the approach described above.

#### Differential gene expression analysis

Visium HD data was pseudobulked across samples and Level 1 annotations using the decoupler package^28^ (v2.1.2). To include only high-quality pseudobulked profiles, we only retained those that had > 10 cells and > 15 total counts. Additionally, to remove artifacts from lowly expressed genes, we performed gene-level filtering in L1 cell types separately to account for intrinsic differences in expression programs across broad cell lineages. We also removed genes that were not present in at least 10% of cells in each pseudobulked profile. We used these pseudobulked profiles for inference with pyDESeq2^85^ (v0.5.4). Reported DE significant genes were obtained by setting the contrast to (early vs advanced)

#### Pathway activity

Pathway activity was estimated using decoupleR^28^ (v2.1.4) and Liana^87^ (v1.7.1) python packages. Cell composition matrix was created for L1 annotations by setting “1” for the cell type each segmented cell was assigned to. Next, for each section **dc.mt.mlm** function was run to use multivariate linear model to estimate the pathway activity.

#### TF-spatial enrichment

To assess the spatial relationship between the L1 cell types and and TF expression, bivariate cosine similarity was calculated using the Liana^87^ (v1.7.1) package. First, for each coronary artery section spatial neighbors were identified using the **spatial_neighbors** function (*bandwidth=200, cutoff=0.1, kernel=’gaussian’, set_diag=True*), and the decoupleR’s ulm function was used to estimate the TF activity in each cell (collecTRI database^75^). Next, the cosine similarity between the cell types and TF-activity in the spatial context was calculated using liana bivariate function and Z-scaled TF expression.

### snATAC processing and analysis in coronary artery

#### Coronary artery sample processing and nuclei isolation

Human coronary artery OCT blocks were cryosectioned to a thickness of 50µm and 15-20 sections were obtained per sample. Single nuclei were isolated from tissue sections using a protocol adapted from 10X Genomics (CG000212) and Grandi et al.^88^. Briefly, OCT-embedded tissue sections were gently dissociated via dounce homogenization and trituration in lysis buffer (10 mM Tris-HCl pH 7.5, 10 mM NaCl, 3 mM MgCl₂, 1% BSA, 0.01% Tween-20, 0.01% NP-40, and 0.001% Digitonin). The suspension was then diluted four-fold in wash buffer (WB; 10 mM Tris-HCl pH 7.5, 10 mM NaCl, 3 mM MgCl₂, 1% BSA, 0.1% Tween-20), filtered through a 30uM cell strainer, and centrifuged at 350g for 5 minutes at 4°C. Pellets were resuspended in WB, and nuclei quality and concentration were assessed via Trypan Blue staining and light microscopy.

Samples with high debris content were further purified using an iodixanol gradient and centrifuged at 3,000g for 20 minutes at 4°C. The fraction located between the 25 and 30% iodixanol gradient was collected and washed in WB, before microscopic re-evaluation. Only samples with high quality nuclei suspension were further processed. In some instances, samples were pooled ensuring they originated from individuals of diverse ancestries. Nuclei were then resuspended in 10X Nuclei Dilution Buffer.

#### snATAC library preparation and sequencing

Subsequent steps of tagmentation and library preparation were performed by the UVA Genome Analysis and Technology Core, RRID:SCR_018883. We used the 10x Genomics Chromium Next GEM Single Cell ATAC Reagent Kits v2 for all snATAC-seq experiments. The full protocols for the snATAC-seq library preparation are available at the following link: https://support.10xgenomics.com/single-cell-atac. snATAC-seq libraries were shipped on dry ice to Psomagen for sequencing on an Illumina NovaSeq X Plus (100 cycles, 2 × 50 bp). Libraries sequenced in-house were run on an Illumina NextSeq 2000 using P2 or P4 100 cycle kits.

#### Histology characterization of coronary samples for snATAC

Adjacent sections of the OCT-embedded tissue used for snATAC-seq were stained with hematoxylin and eosin (H&E), Von Gieson, Oil Red O, and Von Kossa to assess tissue morphology, fibrosis, lipid content, and calcification, respectively. Briefly, coronary artery segments were isolated as described above. Tissues were embedded in OCT blocks, snap-frozen in liquid nitrogen, and stored at −80°C. OCT-embedded human coronary artery segments were cryosectioned at −20°C and 8-μm thickness. A minimum of two sections per sample were placed on each slide and then blindly stained with: Hematoxylin and Eosin (H&E), Von Kossa (VK) and Oil Red O (ORO). Briefly, for H&E, slides were fixed for 10 minutes with 10% Neutral Buffered Formalin, followed by rinsing and incubation with hematoxylin for 5 minutes. This was followed by incubation in bluing solution for 1 minute, washing, incubation in 95% ethanol for 30 seconds and incubation in Eosin-Y for 4 minutes. For VK staining, slides were placed in Neutral Buffered Formalin for 10 minutes, washed, incubated with Silver Nitrate Solution and placed under UV light for 15 minutes. After washing, un-reacted Silver was removed with Sodium Thiosulfate solution for 10 seconds. Slides were washed and counterstained with Nuclear Fast Red solution for 5 minutes. For ORO staining, frozen sections were fixed in 10% Neutral Buffered Formalin solution, washed and stained in Oil Red O solution (Polysciences #s2120) for 15 minutes. After washing, slides were stained in Hematoxylin solution (Richard Allen #7221) for 1 min before rinsing and mounting with aqueous medium. Whole slide images were then captured at approximately 100,000 x 30,000-pixel resolution using a Hamamatsu NanoZoomer S360 Digital Slide Scanner C13220 at the Biorepository and Tissue Research Facility at UVA.

#### Alignment of snATAC libraries

Raw sequencing reads from 10X Genomics unpublished and in-house datasets were aligned to the reference genome using a custom pipeline wrapping Cell Ranger-ATAC (v.2.1.0) (https://www.10xgenomics.com/support/software/cell-ranger-atac/latest) using the hg38 reference genome. The resulting fragment files were used for downstream quality control.

#### Raw snATAC library quality control

The large proportion of low-quality nuclei typically seen in ATAC libraries violates assumptions for filtering cells based on the median absolute deviation (MAD) as done with the scRNA-seq data. Therefore, we opted for an iterative approach that accounted for the heterogeneity (overall quality and sequencing depth) of each library instead. For each individual library, we generated density maps correlating transcription start site (TSS) enrichment vs Number of fragments (N fragments) and used the quadrant with the highest nuclei density to establish a sensible range of values for TSS enrichment (*t*) and N Fragments (*n*). We computed the cartesian product of those iterables to generate (*t* x *n*) possible combinations of parameters. Each combination was fed through a pipeline wrapping snapatac2^89^(v.2.8.0) functions. This workflow was conducted roughly as follows:

1. Retain nuclei with TSS >= *t* and and N fragments >= *n*.
2. Partition the genome into 500bp accessibility bins and select the 250,000 highly variable regions using the **snap.pp.select_features** function setting *max_iter=1*.
3. Remove doublets with the scrublet^90^ implementation within snapatac2 using **snap.pp.scrublet** and **snap.pp.filter_doublets** functions. Nuclei with a doublet ratio of >= 0.5 were removed from downstream analyses.
4. Reduce dimensionality of the data using the snapatac2 spectral algorithm^89^ with the **snap.tl.spectral** function using highly variable accesibility bins.
5. Input the spectral latent space to generate UMAP embeddings with the **snap.tl.umap** function.
6. Generate a kNN graph and perform Leiden clustering using the **snap.pp.knn** and **snap.tl.leiden** functions.

We examined the clustering results and doublet scores of each (*t*, *n*) iteration and selected the combination of parameters that 1) produced the best clustering configuration as determined by the Calinski-Harabasz index computed with scikit-learn (v.1.7.2) 2) yielded the lowest abundance of nuclei with high doublet scores and 3) preserved as many nuclei as possible. The final combinations of *t* and *n* for each library are detailed in **Supplementary Table 10**.

#### Second iteration of QC and Level 1 annotations

We concatenated libraries creating an instance of the **AnnDataSet** class from the snapatac2 ^89^(v.2.8.0) package. Upon merging datasets, we identified 500,000 variable features using the function **snap.pp.select_features** setting the parameter *max_iter=1* and removed ENCODE blacklist regions^91^ using the hg38 v2 BED file obtained from https://github.com/Boyle-Lab/Blacklist. As we observed extensive batch effects across multiple datasets, we performed a first pass of data integration using the harmonypy package ^20^ (v.0.0.10). We used harmony embeddings to generate UMAP embeddings with the **snap.tl.umap** function setting *random_state=0*. A kNN graph was also constructed using Harmony embeddings and then Leiden clustering performed with the function **snap.tl.leiden** at a resolution of 1.0.

We computed gene activity scores, an inferred measure of gene expression based on accessibility within a gene promoter/body using snapatac2 **snap.pp.make_gene_matrix** function. Gene activity scores were normalized using the scanpy^71^ **normalize_total** and **log1p** functions as described above. To smooth the inferred signal, we used the scanpy **sc.external.pp.magic** function to impute gene activity scores setting *solver=”approximate”*. We used markers from the MetaPlaq RNA dataset in addition to gene score cluster markers from Wilcoxon rank-sum tests to conduct a first pass of annotations. During this round of level 1 (L1) annotations, we found a cluster characterized by mixed-lineage marker gene activity and elevated scrublet^90^ scores, likely representing remaining doublets. This low-quality cluster was removed for downstream analyses.

#### Integration Benchmarking

Upon removing remaining low-quality nuclei from the concatenated AnnDataSet, we conducted a benchmark to evaluate the performance of linear embeddings and deep learning models as described in the above section. We consider this an important part of this study as benchmarking of batch correction methods has been solely carried out for scRNA-seq datasets. Furthermore, the number of available methods for this task is not nearly as large as that of single-cell transcriptomics data. Therefore, the cardiovascular field still lacks a frame of reference regarding the performance of harmonization algorithms for single-cell epigenomics data. The address this gap, the following integration algorithms were assessed using baseline spectral embeddings as a control:

### Harmony

To perform batch correction with Harmony^20^ (v.0.0.10), we used the concatenated AnnDataset as input to the snapatac2 wrapper function **snap.pp.harmony** setting the parameter *max_iter_harmony=20*. Harmony embeddings were used for downstream clustering with the functions **snap.pp.knn** and **snap.tl.leiden** with default parameters.

### PeakVI

PeakVI^41^ is a probabilistic framework included in the scVI suite designed for the analysis of scATAC-seq data that uses a variational autoencoder (VAE) architecture to learn a low-dimensional latent representation of chromatin accessibility profiles. Batch correction is performed by conditioning the decoder on batch identity during the model training process, allowing the latent space to capture biologically meaningful variation while reducing technical batch effects. To use this framework, the concatenated object was subset to the 500,000 highly variable features found as previously described and then to peaks present in more than 5% of cells. With *scvi.settings.seed=420*, the setup before model creation and training was performed using the **scvi.model.PEAKVI.setup_anndata** from the scvitools package^26^ (v.1.1.5), where the batch key was provided. The model was instantiated using the class **scvi.model.PEAKVI** and then trained with the following parameters (*n_layers=2, n_hidden=sqrt(number of regions), n_latent=sqrt(n_hidden), dropout_rate=0.1, model_depth=True, latent_distribution=’normal’*) to generate the batch corrected embeddings. The embedding was extracted using the **get_latent_representation()** method and saved. UMAP embeddings based on this embedding were obtained using the **scanpy.pp.neighbors** and **scanpy.tl.umap** (*min_dist=0.1*) functions.

### Graph-Linked Unified Embedding (GLUE)

GLUE^40^ jointly trains omics-specific variational autoencoders (VAEs) and leverages a biologically informed guidance graph to simultaneously generate within-modality embeddings and cross-modality alignment in a shared latent space. To perform batch correction, batch labels are incorporated as covariates in the omics-specific VAEs. The VAE can be trained in a supervised or unsupervised mode. In the supervised mode, provided cell annotations are used to guide cross-modality alignment by encouraging cells with shared labels across modalities to occupy similar positions in the shared latent space during joint optimization. The batch correction of the embeddings produced by both modes were evaluated. Before the VAEs were jointly trained, the guidance graph was produced. The genomic coordinates of the scRNA-seq features were annotated using the **scglue.data.get_gene_annotation** function from the scglue package^40^ (v.0.4.0) by supplying the GTF annotation used for scRNA-seq read alignment (Cell Ranger GRCh38 - 2024A). Afterwards, the guidance graph was constructed using the **rna_anchored_guidance_graph** function by supplying the annotated scRNA-seq and snATAC-seq data. The graph was checked using the **graph.check_graph** function. Then, the VAEs were jointly trained to produce the snATAC-seq embedding. First the datasets wre configured using the the **scglue.models.configure_dataset** function (*NB* distribution, and keys provided for highly variable genes, raw count matrices, low dimensional embeddings, and sample annotations). The modality-specific embeddings were produced using the function **scglue.models.fit_SCGLUE** using default parameters (48 epochs, 4 patience). The co-embedding of the scRNA and snATAC was also performed in the supervised mode, using the above function with the same parameters, in addition to curated cell L1 annotations from the RNA reference.

#### Refinement of L1 annotations

Upon coembedding RNA and ATAC modalities, GLUE label transfer was then performed to validate annotated L1 cell classes within the snATAC data. We used the **scglue.data.transfer_labels** function with default parameters providing L1 scRNA annotations.

We investigated prediction confidence scores, and the correspondence of GLUE transferred labels with our manual annotations to come up with a final set of L1 ATAC annotations. Overall, we observed high prediction confidence (GLUE-computed confidence score > 0.8) for major L1 cell types including SMCs, ECs, macrophages, fibroblasts, B cells, and pericytes with an overall confidence score of 0.98. A small cluster that was manually labeled as SMCs in the was reannotated as pericytes, and mast cells refined to plasma cells based on their strong immunoglobulin gene activity scores and accessibility profiles. Labels transferred as neurons were relabeled as unknown as we did not detect marked accessibility at the TSS of canonical neuron markers identified in the scRNA-seq data.

#### ATAC level 2 (L2) annotations and murine gene activity enrichment

L1 compartments for relevant cells were subset to a single major cell type of interest (e.g., SMC and Endothelial cells). For each subset, we conducted a new round of feature selection using snapatac2 **snap.pp.select_features** function to extract 250,000 highly variable regions. These regions were used as input to the **snap.tl.spectral** function to obtain a low-dimensional representation of the target cell type. We then used PeakVI^41^ to perform a new round of batch correction in the data subset by creating an instance of the class **scvi.model.PEAKVI** and setting *n_layers_encoder=2* and *n_layers_decoder=2*.

To add level 2 (L2) annotations, we performed Leiden clustering across a sensible range of resolutions and used gene activity scores of canonical subtype markers identified within our scRNA-seq reference. We further validated SMC ATAC annotations using murine ligea-traced SMC signatures ^10^ with decoupler^28^ (v.2.1.1) as previously done with scRNA-seq clusters.

#### Peak calling and differential accessibility analysis

Peaks were called using the **snap.tl.macs3** function from snpatac2^89^ (v.2.8.0) (*qvalue=0.01, shift=-75, extsize=150, nolambda=False*). During peak calling, the ENCODE blacklist BED file ^91^ was provided to filter problematic regions. Next, the **snap.tl.merge_peaks** function was used to produce a unified, non-overlapping, and fixed-width 500 bp peaks using default parameters and the hg38 genome reference provided by snapatac2. The **snap.pp.make_peak_matrix** function was used to generate a cell-by-peak matrix for downstream analyses. Differentially accessible regions (DARs) were then detected for each of the L1 and L2 cell classes using the **scanpy.tl.rank_genes_groups function** from scanpy^71^ (v.1.11.4) with parameters (*method=”wilcoxon”, corr_method=”benjamini-hochberg”*). The output peaks were then filtered (FDR < 0.01, Log2FC > 1). The genomic region enrichment analysis tool (GREAT)^92^(v.4.0.4) was then used to identify functional terms enriched within each set of L1 and L2 DARs. DARs characterization, including distribution within chromosomes, neighboring region distances and overlap with ENCODE bulk profiles was performed with our in-house GenomicDistributions R package^93^ (v.1.10.0).

#### Motif analyses

Pseudobulked motif enrichment analysis was performed on L1 and L2 DARs using HOMER^94^ (v.5.1). Known and de novo motifs were identified against the hg38 genome (*-size 500*).

Furthermore, to identify highly accessible motifs in a per-cell basis, we also used the pychromvar package (v.0.0.4), a Python implementation of the original chromVar algorithm^95^ Roughly, we created a peak anndata file as described above. Peak sequences were extracted using the **add_peak_seq** function. We then estimated GC bias and generated a set of background peaks using the **add_gc_bias** and **get_bg_peaks.** Motifs were extracted using the function **fetch_motifs** with the pyjaspar (v3.0.0) Python package using the JASPAR2024 release and subset to contain only human motifs. Motif deviations were computed using the **match_motifs** and **compute_deviations** functions. Marker motifs were computed using a Wilcoxon Rank Sum Test with the **scanpy.tl.rank_genes_groups** function.

#### Enhancer-to-gene link predictions

The scE2G pipeline ^42^ is a supervised classifier adapted for snATAC and multiome data from ENCOD-rE2G. It integrates multiple features, including a pseudobulked ABC score, where 3D contact is estimated by an inverse function of genomic distance among others^96^. In each cell type, scE2G scores every candidate element-gene pair (candidate elements are ATAC peaks called with MACS2^97^ within 5mb of the TSS of the gene) by integrating several features, such as ABC scores, ATAC signal at a given promoter and gene density. To run the pipeline, we created pseudobulked fragment files for each L1 and L2 annotation using the **snap.pp.export_fragments** function from snapATAC2. These fragment files were preprocessed for the scE2G pipeline using a custom NextFlow pipeline that sorted each fragment, bgzip compressed and then indexed them with tabix setting the parameter *–preset bed*.

Next, we used the scE2G (scATAC_powerlaw_v3) model (v.1.3) to predict enhancer-gene interactions in L1 and L2 pseudobulked fragment files. A configuration file pointing to preprocessed fragment files was used as input to run the snakemake pipeline for each cell type separately. Key parameters in the config included *max_cell_count=40000*, *fragments_preprocessed=True* and *benchmark_performance=False*. Regulatory enhancer-gene interactions were defined as element-gene pairs with a score greater than 0.171. The score threshold of 0.171 was determined as the score yielding 70% recall when evaluating predictions in K562 cells against CRISPRi-validated enhancer-gene pairs. Bigwig files generated from the pipeline were used for visualization in the UCSC genome browser. BEDPE files with the peak-to gene links were converted to bigInteract files using an in-house NextFlow pipeline.

For quantifying the degree of specificity for EC enhancer-gene connections, we computed the Shannon entropy, and subsequently specificity, for each gene across L2 annotations as follows:

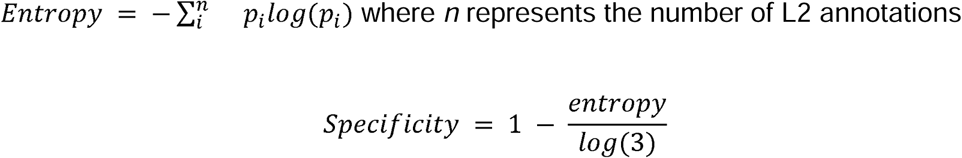

#### Enhancer matrix factorization and genomic region overlaps

To perform a global comparison of the enhancer landscape of L1 and L2 annotations, we used two orthogonal approaches. In the first approach, we created a matrix where rows are L1 or L2 annotations and columns depict genes. Each value in the matrix represents the number of enhancer connections to a gene in a given cell annotation. We normalized raw enhancer counts by total number of enhancer links in a given cell annotation and then log-transformed normalized counts with the numpy function **log1p.** Prior to dimensionality reduction, we computed the variance of log counts, computed the median of the variance and removed genes falling under that threshold. High-variance normalized counts were standardized with Z-score scaling using the **StandardScaler** class from the scikit-learn package (v.1.7.2). Singular value decomposition (SVD) was then run using the **PCA** class of the scikit-learn package.

In the second approach, we loaded filtered enhancer predictions using a custom wrapper function in R (v.4.3.1) and removed duplicated enhancers. We performed region overlaps taking the cartesian product of L1 annotations to do an all-vs-all comparison. We counted overlaps using the **countOverlaps** function from the GenomicRanges package (v.1.54.1) and computed the proportion between two different region sets as follows:

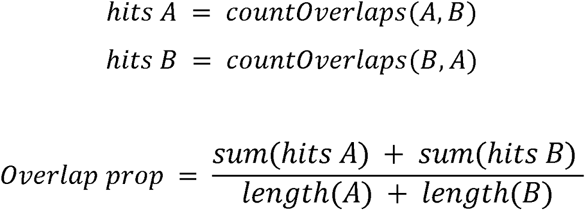

#### LD Score regression

Reference files, including LD scores fom the baseline model (v1.2) and hapmap (hm3) reference regression weights were extracted from the following Zenodo link: https://zenodo.org/records/10515792. GWAS summary statistic files were processed using our in-house cojo package as described above. Next, we used a custom NextFlow pipeline wrapping the **munge_sumstats.py** from the ldsc Github repository (https://github.com/bulik/ldsc) to convert these summary files to a LDSC-compatible format.

Prior to LDSC regression, we wrote a NextFlow pipeline to preprocess DARs. DAR coordinates were first lifted over from hg38 to hg19 using the UCSC binary liftover and corresponding chain files. We then used these hg19 BED files to create annotation files across 22 chromosomes for each L1/L2 annotation. LD scores were then computed for each of these annotations. Finally, we implemented a new pipeline to create input .ldcts files using the custom **create_ldcts_file.py** script. Finally, LDSC regression was performed using the proper cell-type-specific analysis parameters and munged summary stats as described in https://github.com/bulik/ldsc/wiki/Cell-type-specific-analyses.

### eGRN analysis using SCENIC+

#### Data preprocessing

SCENIC+^54^**(v.1.0a2)** was used to identify regulons for L1-defined major cell types and L2-defined SMC and EC subtypes. To increase robustness of the analysis, scRNA-seq and snATAC-seq references were subsetted to overlapping cell-type annotations. The resulting cell-by-gene and cell-by-peak matrices were used as inputs. Candidate regions for TF binding and *cis*-regulatory topics were identified using pycisTopic ^98^(**v.2.0a0**), followed by motif enrichment analysis using pycistarget (**v.1.1**).

#### eGRN construction

Pseudo-multiome metacells were generated by sampling cells across modalities within each cell type. TF-gene and region-gene links were inferred using GRNBoost^99^ (**arboreto v.0.1.6**), and regulons for each TF were defined by motif-enriched regions and their linked genes. Regulon activity was quantified per cell using AUCell^100^ (**ctxcore v.0.2.0**) based on ranked gene expression and accessibility profiles. Regulon specificity scores (RSS) were calculated using the gene-based and region-based AUC scores. Networks for selected TFs were generated and plotted by aggregating individual eRegulons containing the target TF and target genes were shown based on cell-specific pseudobulked L1 differential expression in advanced lesions or effector gene status. Genomic tracks of chromatin accessibility in putative enhancer regions within select eRegulons were also inspected for cellular subtype-specific activity. All GRNs predicted for each set of cellular annotations can be found in **Supplementary Table 13**.

#### In silico perturbations

A knockout simulation for BNC2 was performed by setting its inferred activity to zero and propagating the effect through the learned TF-target weight matrix computed at the SMC L2 annotation level. Changes in predicted gene expression levels were evaluated by comparing expression to their original, unperturbed baseline. These changes were characterized at the gene level. Additionally, the impact of these changes on cellular identity was assessed by projecting the velocity of perturbed profiles onto a PCA embedding of baseline regulon activity.

#### L1 regulon module identification and functional enrichment

Pairwise Pearson R correlations were computed on region-based AUC scores among activating regulons using the pandas (**v.2.1.1**) *.corr()* method. SCENIC+ metacells were first filtered to the cell type of interest (e.g. SMC or EC). The correlation was then calculated using the region-based AUC scores from each metacell for each regulon in the pairwise comparison. Hierarchical clustering was then performed on the resulting correlation matrix using the Seaborn (**v.0.13.2**) *sns.clustermap* function. Dendrogram cuts were selected based on cophenetic distance to yield 5-7 modules per cell type. GREAT^92^ (**v.4.0.4**) functional enrichment was then performed on the union of regions between the top 10 regulons per module ranked by region-based RSS. Each term was ranked by -Log(FDR) and the top 5 terms per module were shown (P *adj* < 0.05).

#### Gene set enrichment of individual L1 regulons

GSEA was performed with the gseapy^101^(**v.1.1.12**) package using the *gp.enrich* function. First, DEGs upregulated in advanced lesions in SMCs and ECs were identified (*P adj* < 0.05, |Log_2_FC| > 1). Effector genes for CAD in SMCs and ECs were also compiled as an orthogonal set for gene set enrichment. The background used to perform the enrichment was all gene targets in any regulon predicted by SCENIC+. The upregulated DEG gene set and CAD effector gene sets were separately tested for enrichment in each individual regulon. The -Log_10_(*P adj*) for DEG enrichment was plotted against the -Log_10_(*P adj*) for effector gene enrichment.

#### CAD MVP credible set processing and locus visualization

The credible set was extracted from the corresponding publication^7^ and converted into a GRanges object using the GenomicRanges package (v.1.54.1). Then, rsIDs were extracted with the function **snpsByOverlaps** function from the BSgenome R package (v.1.70.2) and the SNPlocs.Hsapiens.dbSNP155.GRCh37 package. The hg38 coordinates of the obtained rsIDs were then retrieved with the function **snpsById** and the SNPlocs.Hsapiens.dbSNP155.GRCh38 R package. Prior to overlaps, 100 bp windows were created around the center of the SNP hg38 coordinates (50 bp upstream/dowstream from the SNP location). Windows were then annotated with genomic partitions using the ChiPSeekR package^102^ (v1.38.0) setting the partitions priority in the following order (Promoter, 3UTR, 5UTR, Exon, Intron, Downstream and Intergenic) and defining TSS coordinates as (-50, 2000). Windows were then overlapped with DARs, scE2G predicted enhancers and H3K27ac regions using custom functions wrapping GenomicRanges package (v.1.54.1). For generation of LocusZoom plots, summary statistics from CAD^7^ were munged to retrieve hg38 coordinates using the MungeSumStats R package^92,103^ (v.1.10.1). Loci of interest were then visualized using the locuszoomr R package^104^ (v.0.3.8) using a flank distance of 1e5 and annotations from the EnsDb.Hsapiens.v86 package.

#### TF binding predictions

To obtain predictions of TF motifs disrupted by credible set SNPs in L1 DARs, we used the motifbreakR R package^105^ (v2.16.0) using a motif superset with the following databases: HOCOMOCO v11/v13, JASPAR Core and JASPAR 2022.

## Data availability

### Transcriptomics

Publicly available scRNA-seq datasets were obtained from the Gene Expression Omnibus (GEO) and the corresponding accession numbers are listed in Supplemental Table 1. Raw and processed bulk RNA-sequencing data is publicly available in GEO under the accession number GSE225650. Visium HD data from the COSMOS cohort will be made available upon publication.

### Epigenomics

Human carotid and femoral artery plaque snATAC fragment files were directly obtained from the authors of the study^17^. Tibial artery snATAC data from ENCODE is available through GEO (GSE286599). In-house coronary artery snATAC-seq data from our previous publication^8^ is available on the GEO database (GSE175621 and GSE188422) Newly generated snATAC data from human coronary arteries from the COSMOS cohort will be made available upon publication.

## Code availability

All scripts used in this study will be deposited on our GitHub page: https://github.com/MillerLab-CPHG

## Notes

### Author Declarations

Ethics committee/Institutional Review Board of Stanford University gave ethical approval for this work. Ethics committee/Institutional Review Board of the University of Virginia gave ethical approval for this work.

## References

1. Global Burden of Cardiovascular Diseases and Risks 2023 Collaborators. Global, regional, and national burden of cardiovascular diseases and risk factors in 204 countries and territories, 1990-2023. J. Am. Coll. Cardiol. 86, 2167–2243 (2025).

2. Björkegren, J. L. M. & Lusis, A. J. Atherosclerosis: Recent developments. Cell 185, 1630–1645 (2022).

3. Matsuura, Y., Kanter, J. E. & Bornfeldt, K. E. Highlighting residual atherosclerotic cardiovascular disease risk. Arterioscler. Thromb. Vasc. Biol. 39, e1–e9 (2019).

4. Evrard, S. M. et al. Endothelial to mesenchymal transition is common in atherosclerotic lesions and is associated with plaque instability. Nat. Commun. 7, 11853 (2016).

5. Liu, M. & Gomez, D. Smooth muscle cell phenotypic diversity. Arterioscler. Thromb. Vasc. Biol. 39, 1715–1723 (2019).

6. Aragam, K. G. et al. Discovery and systematic characterization of risk variants and genes for coronary artery disease in over a million participants. Nat. Genet. 54, 1803–1815 (2022).

7. Tcheandjieu, C. et al. Large-scale genome-wide association study of coronary artery disease in genetically diverse populations. Nat Med 28, 1679–1692 (2022).

8. Turner, A. W. et al. Single-nucleus chromatin accessibility profiling highlights regulatory mechanisms of coronary artery disease risk. Nat Genet 54, 804–816 (2022).

9. Mosquera, J. V. et al. Integrative single-cell meta-analysis reveals disease-relevant vascular cell states and markers in human atherosclerosis. Cell Rep. 42, 113380 (2023).

10. Li, D. Y. et al. Vascular smooth muscle cell state trajectories mediate molecular mechanisms of coronary disease risk. Nat. Commun. (2026) doi:10.1038/s41467-026-70530-z.

11. Bleckwehl, T. et al. Encompassing view of spatial and single-cell RNA sequencing renews the role of the microvasculature in human atherosclerosis. Nat Cardiovasc Res 4, 26–44 (2025).

12. Traeuble, K. et al. Integrated single-cell atlas of human atherosclerotic plaques. Nat Commun 16, 8255 (2025).

13. Pauli, J. et al. Single cell spatial transcriptomics integration deciphers the morphological heterogeneity of atherosclerotic carotid arteries. Nat Commun 16, 11282 (2025).

14. Amrute, J. M. et al. Targeting modulated vascular smooth muscle cells in atherosclerosis via FAP-directed immunotherapy. Science eadx1736 (2026).

15. Maurano, M. T. et al. Systematic localization of common disease-associated variation in regulatory DNA. Science 337, 1190–1195 (2012).

16. Buenrostro, J. D., Giresi, P. G., Zaba, L. C., Chang, H. Y. & Greenleaf, W. J. Transposition of native chromatin for fast and sensitive epigenomic profiling of open chromatin, DNA-binding proteins and nucleosome position. Nat Methods 10, 1213–1218 (2013).

17. Örd, T. et al. Single-Cell Epigenomics and Functional Fine-Mapping of Atherosclerosis GWAS Loci. Circ Res 129, 240–258 (2021).

18. Lotfollahi, M. et al. Mapping single-cell data to reference atlases by transfer learning. Nat. Biotechnol. 40, 121–130 (2022).

19. Stuart, T. et al. Comprehensive Integration of Single-Cell Data. Cell 177, 1888–1902.e21 (2019).

20. Korsunsky, I. et al. Fast, sensitive and accurate integration of single-cell data with Harmony. Nat Methods 16, 1289–1296 (2019).

21. Hie, B., Bryson, B. & Berger, B. Efficient integration of heterogeneous single-cell transcriptomes using Scanorama. Nat Biotechnol 37, 685–691 (2019).

22. Andreatta, M. & Carmona, S. J. STACAS: Sub-Type Anchor Correction for Alignment in Seurat to integrate single-cell RNA-seq data. Bioinformatics 37, 882–884 (2021).

23. de Leeuw, C. A., Mooij, J. M., Heskes, T. & Posthuma, D. MAGMA: generalized gene-set analysis of GWAS data. PLoS Comput Biol 11, e1004219 (2015).

24. Zhang, M. J. et al. Polygenic enrichment distinguishes disease associations of individual cells in single-cell RNA-seq data. Nat. Genet. 54, 1572–1580 (2022).

25. Polański, K. et al. Bin2cell reconstructs cells from high resolution Visium HD data. Bioinformatics 40, (2024).

26. Lopez, R., Regier, J., Cole, M. B., Jordan, M. I. & Yosef, N. Deep generative modeling for single-cell transcriptomics. Nat Methods 15, 1053–1058 (2018).

27. Timshel, P. N., Thompson, J. J. & Pers, T. H. Genetic mapping of etiologic brain cell types for obesity. Elife 9, (2020).

28. Badia-I-Mompel, P., et al. decoupleR: ensemble of computational methods to infer biological activities from omics data. Bioinform. Adv. 2, vbac016 (2022).

29. Caus, M. et al. Vitamin D Receptor From VSMCs Regulates Vascular Calcification During CKD: A Potential Role for miR-145a. Arterioscler Thromb Vasc Biol 43, 1533–1548 (2023).

30. Fessler, M. B. Liver X Receptor: Crosstalk Node for the Signaling of Lipid Metabolism, Carbohydrate Metabolism, and Innate Immunity. Curr Signal Transduct Ther 3, 75–81 (2008).

31. Majdalawieh, A., Zhang, L., Fuki, I. V., Rader, D. J. & Ro, H.-S. Adipocyte enhancer-binding protein 1 is a potential novel atherogenic factor involved in macrophage cholesterol homeostasis and inflammation. Proc Natl Acad Sci U S A 103, 2346–2351 (2006).

32. Pruenster, M. et al. The Duffy antigen receptor for chemokines transports chemokines and supports their promigratory activity. Nat Immunol 10, 101–108 (2009).

33. Guo, X. et al. Endothelial ACKR1 is induced by neutrophil contact and down-regulated by secretion in extracellular vesicles. Front Immunol 14, 1181016 (2023).

34. Schnitzler, G. R. et al. Convergence of coronary artery disease genes onto endothelial cell programs. Nature 626, 799–807 (2024).

35. Stolze, L. K. et al. Systems Genetics in Human Endothelial Cells Identifies Non-coding Variants Modifying Enhancers, Expression, and Complex Disease Traits. Am J Hum Genet 106, 748–763 (2020).

36. Love, M. I., Huber, W. & Anders, S. Moderated estimation of fold change and dispersion for RNA-seq data with DESeq2. Genome Biol 15, 550 (2014).

37. Schubert, M. et al. Perturbation-response genes reveal signaling footprints in cancer gene expression. Nat Commun 9, 20 (2018).

38. Kavousi, M. et al. Multi-ancestry genome-wide study identifies effector genes and druggable pathways for coronary artery calcification. Nat Genet 55, 1651–1664 (2023).

39. Mueller, P. A. et al. Coronary Artery Disease Risk-Associated Gene and Its Product Lipid Phosphate Phosphatase 3 Regulate Experimental Atherosclerosis. Arterioscler Thromb Vasc Biol 39, 2261–2272 (2019).

40. Cao, Z.-J. & Gao, G. Multi-omics single-cell data integration and regulatory inference with graph-linked embedding. Nat Biotechnol 40, 1458–1466 (2022).

41. Ashuach, T., Reidenbach, D. A., Gayoso, A. & Yosef, N. PeakVI: A deep generative model for single-cell chromatin accessibility analysis. Cell Rep Methods 2, 100182 (2022).

42. Sheth, M. U., et al. Mapping enhancer-gene regulatory interactions from single-cell data. bioRxiv (2024) doi:10.1101/2024.11.23.624931.

43. Finucane, H. K. et al. Heritability enrichment of specifically expressed genes identifies disease-relevant tissues and cell types. Nat Genet 50, 621–629 (2018).

44. Giri, A. et al. Trans-ethnic association study of blood pressure determinants in over 750,000 individuals. Nat Genet 51, 51–62 (2019).

45. Hartiala, J. A. et al. Genome-wide analysis identifies novel susceptibility loci for myocardial infarction. Eur Heart J 42, 919–933 (2021).

46. Huang, X. et al. Critical role for the Ets transcription factor ELF-1 in the development of tumor angiogenesis. Blood 107, 3153–3160 (2006).

47. Zhuang, T. et al. Cell-Specific Effects of GATA (GATA Zinc Finger Transcription Factor Family)-6 in Vascular Smooth Muscle and Endothelial Cells on Vascular Injury Neointimal Formation. Arterioscler Thromb Vasc Biol 39, 888–901 (2019).

48. Jeong, K. et al. Nuclear Focal Adhesion Kinase Controls Vascular Smooth Muscle Cell Proliferation and Neointimal Hyperplasia Through GATA4-Mediated Cyclin D1 Transcription. Circ Res 125, 152–166 (2019).

49. Rykaczewska, U. et al. Abstract 308: Foxc1 is a major transcription factor influencing smooth muscle cell activation in atherosclerotic plaques. Arterioscler. Thromb. Vasc. Biol. 39, (2019).

50. Marcelo, K. L., Goldie, L. C. & Hirschi, K. K. Regulation of endothelial cell differentiation and specification. Circ Res 112, 1272–1287 (2013).

51. de Vries, P. et al. A multi-trait genome-wide association study of coronary artery disease and subclinical atherosclerosis traits. Res Sq (2025) doi:10.21203/rs.3.rs-6456056/v1.

52. Tan, J. M. E. et al. PRDM16 regulates smooth muscle cell identity and atherosclerotic plaque composition. Nat Cardiovasc Res 4, 1573–1588 (2025).

53. Wong, D. et al. FHL5 Controls Vascular Disease-Associated Gene Programs in Smooth Muscle Cells. Circ Res 132, 1144–1161 (2023).

54. Bravo González-Blas, C., et al. SCENIC+: single-cell multiomic inference of enhancers and gene regulatory networks. Nat Methods 20, 1355–1367 (2023).

55. Chen, Y. et al. IRF6 induces endothelial dysfunction through the transcriptional activation of NDRG1 and aggravates low shear stress-mediated atherosclerosis. iScience 29, 115127 (2026).

56. Xue, Q., Wang, X., Deng, X., Huang, Y. & Tian, W. CEMIP regulates the proliferation and migration of vascular smooth muscle cells in atherosclerosis through the WNT-beta-catenin signaling pathway. Biochem. Cell Biol. 98, 249–257 (2020).

57. Hernández-García, R., Iruela-Arispe, M. L., Reyes-Cruz, G. & Vázquez-Prado, J. Endothelial RhoGEFs: A systematic analysis of their expression profiles in VEGF-stimulated and tumor endothelial cells. Vascul Pharmacol 74, 60–72 (2015).

58. Maiwald, S. et al. A rare variant in MCF2L identified using exclusion linkage in a pedigree with premature atherosclerosis. Eur J Hum Genet 24, 86–91 (2016).

59. Wang, L. et al. Peakwide mapping on chromosome 3q13 identifies the kalirin gene as a novel candidate gene for coronary artery disease. Am J Hum Genet 80, 650–663 (2007).

60. Mori, M. et al. CD163 Macrophages Induce Endothelial-to-Mesenchymal Transition in Atheroma. Circ Res 135, e4–e23 (2024).

61. Craig, M. P. et al. Etv2 and fli1b function together as key regulators of vasculogenesis and angiogenesis. Arterioscler Thromb Vasc Biol 35, 865–876 (2015).

62. Cario-Toumaniantz, C. et al. RhoA guanine exchange factor expression profile in arteries: evidence for a Rho kinase-dependent negative feedback in angiotensin II-dependent hypertension. Am J Physiol Cell Physiol 302, C1394–404 (2012).

63. Hodonsky, C. J. et al. Multi-ancestry genetic analysis of gene regulation in coronary arteries prioritizes disease risk loci. Cell Genom 4, 100465 (2024).

64. Depuydt, M. A. C. et al. Microanatomy of the Human Atherosclerotic Plaque by Single-Cell Transcriptomics. Circ Res 127, 1437–1455 (2020).

65. Mocci, G. et al. Single-Cell Gene-Regulatory Networks of Advanced Symptomatic Atherosclerosis. Circ Res 134, 1405–1423 (2024).

66. Dobin, A. et al. STAR: ultrafast universal RNA-seq aligner. Bioinformatics 29, 15–21 (2013).

67. Germain, P.-L., Lun, A., Garcia Meixide, C., Macnair, W. & Robinson, M. D. Doublet identification in single-cell sequencing data using. F1000Res **10**, 979 (2021).

68. Yang, S. et al. Decontamination of ambient RNA in single-cell RNA-seq with DecontX. Genome Biol 21, 57 (2020).

69. McCarthy, D. J., Campbell, K. R., Lun, A. T. L. & Wills, Q. F. Scater: pre-processing, quality control, normalization and visualization of single-cell RNA-seq data in R. Bioinformatics 33, 1179–1186 (2017).

70. Hafemeister, C. & Satija, R. Normalization and variance stabilization of single-cell RNA-seq data using regularized negative binomial regression. Genome Biol 20, 296 (2019).

71. Wolf, F. A., Angerer, P. & Theis, F. J. SCANPY: large-scale single-cell gene expression data analysis. Genome Biol 19, 15 (2018).

72. Luecken, M. D. et al. Benchmarking atlas-level data integration in single-cell genomics. Nat Methods 19, 41–50 (2022).

73. Butler, A., Hoffman, P., Smibert, P., Papalexi, E. & Satija, R. Integrating single-cell transcriptomic data across different conditions, technologies, and species. Nat. Biotechnol. 36, 411–420 (2018).

74. Xu, C. et al. Probabilistic harmonization and annotation of single-cell transcriptomics data with deep generative models. Mol Syst Biol 17, e9620 (2021).

75. Müller-Dott, S. et al. Expanding the coverage of regulons from high-confidence prior knowledge for accurate estimation of transcription factor activities. Nucleic Acids Res 51, 10934–10949 (2023).

76. Franceschini, N. et al. GWAS and colocalization analyses implicate carotid intima-media thickness and carotid plaque loci in cardiovascular outcomes. Nat Commun 9, 5141 (2018).

77. Mishra, A. et al. Stroke genetics informs drug discovery and risk prediction across ancestries. Nature 611, 115–123 (2022).

78. Jansen, I. E. et al. Genome-wide meta-analysis identifies new loci and functional pathways influencing Alzheimer’s disease risk. Nat Genet 51, 404–413 (2019).

79. Sudlow, C. et al. UK biobank: an open access resource for identifying the causes of a wide range of complex diseases of middle and old age. PLoS Med 12, e1001779 (2015).

80. He, Y., Koido, M., Shimmori, Y. & Kamatani, Y. GWASLab: a Python package for processing and visualizing GWAS summary statistics. (2023) doi:10.51094/jxiv.370.

81. Slenders, L. et al. Intersecting single-cell transcriptomics and genome-wide association studies identifies crucial cell populations and candidate genes for atherosclerosis. Eur Heart J Open 2, oeab043 (2022).

82. Finucane, H. K. et al. Partitioning heritability by functional annotation using genome-wide association summary statistics. Nat Genet 47, 1228–1235 (2015).

83. Zeisel, A. et al. Molecular Architecture of the Mouse Nervous System. Cell 174, 999–1014.e22 (2018).

84. Skene, N. G. et al. Genetic identification of brain cell types underlying schizophrenia. Nat Genet 50, 825–833 (2018).

85. Muzellec, B., Teleńczuk, M., Cabeli, V. & Andreux, M. PyDESeq2: a python package for bulk RNA-seq differential expression analysis. Bioinformatics 39, (2023).

86. van de Geijn, B., McVicker, G., Gilad, Y. & Pritchard, J. K. WASP: allele-specific software for robust molecular quantitative trait locus discovery. Nat Methods 12, 1061–1063 (2015).

87. Dimitrov, D. et al. LIANA+ provides an all-in-one framework for cell-cell communication inference. Nat Cell Biol 26, 1613–1622 (2024).

88. Grandi, F. C., Modi, H., Kampman, L. & Corces, M. R. Chromatin accessibility profiling by ATAC-seq. Nat Protoc 17, 1518–1552 (2022).

89. Zhang, K., Zemke, N. R., Armand, E. J. & Ren, B. A fast, scalable and versatile tool for analysis of single-cell omics data. Nat Methods 21, 217–227 (2024).

90. Wolock, S. L., Lopez, R. & Klein, A. M. Scrublet: Computational Identification of Cell Doublets in Single-Cell Transcriptomic Data. Cell Syst 8, 281–291.e9 (2019).

91. Amemiya, H. M., Kundaje, A. & Boyle, A. P. The ENCODE Blacklist: Identification of Problematic Regions of the Genome. Sci Rep 9, 9354 (2019).

92. McLean, C. Y. et al. GREAT improves functional interpretation of cis-regulatory regions. Nat Biotechnol 28, 495–501 (2010).

93. Kupkova, K. et al. GenomicDistributions: fast analysis of genomic intervals with Bioconductor. BMC Genomics 23, 299 (2022).

94. Zhang, F. & Chen, J. Y. HOMER: a human organ-specific molecular electronic repository. BMC Bioinformatics 12 Suppl 10, S4 (2011).

95. Schep, A. N., Wu, B., Buenrostro, J. D. & Greenleaf, W. J. chromVAR: inferring transcription-factor-associated accessibility from single-cell epigenomic data. Nat Methods 14, 975–978 (2017).

96. Fulco, C. P. et al. Activity-by-contact model of enhancer-promoter regulation from thousands of CRISPR perturbations. Nat Genet 51, 1664–1669 (2019).

97. Feng, J., Liu, T., Qin, B., Zhang, Y. & Liu, X. S. Identifying ChIP-seq enrichment using MACS. Nat Protoc 7, 1728–1740 (2012).

98. Bravo González-Blas, C., et al. cisTopic: cis-regulatory topic modeling on single-cell ATAC-seq data. Nat Methods 16, 397–400 (2019).

99. Moerman, T. et al. GRNBoost2 and Arboreto: efficient and scalable inference of gene regulatory networks. Bioinformatics 35, 2159–2161 (2019).

100. Aibar, S. et al. SCENIC: single-cell regulatory network inference and clustering. Nat Methods 14, 1083–1086 (2017).

101. Fang, Z., Liu, X. & Peltz, G. GSEApy: a comprehensive package for performing gene set enrichment analysis in Python. Bioinformatics 39, (2023).

102. Yu, G., Wang, L.-G. & He, Q.-Y. ChIPseeker: an R/Bioconductor package for ChIP peak annotation, comparison and visualization. Bioinformatics 31, 2382–2383 (2015).

103. Murphy, A. E., Schilder, B. M. & Skene, N. G. MungeSumstats: a Bioconductor package for the standardization and quality control of many GWAS summary statistics. Bioinformatics 37, 4593–4596 (2021).

104. Lewis, M. J. & Wang, S. locuszoomr: an R package for visualizing publication-ready regional gene locus plots. Bioinform Adv 5, vbaf006 (2025).

105. Coetzee, S. G., Coetzee, G. A. & Hazelett, D. J. motifbreakR: an R/Bioconductor package for predicting variant effects at transcription factor binding sites. Bioinformatics 31, 3847–3849 (2015).

